# Serological signatures of SARS-CoV-2 infection: Implications for antibody-based diagnostics

**DOI:** 10.1101/2020.05.07.20093963

**Authors:** Jason Rosado, Stéphane Pelleau, Charlotte Cockram, Sarah Hélène Merkling, Narimane Nekkab, Caroline Demeret, Annalisa Meola, Solen Kerneis, Benjamin Terrier, Samira Fafi-Kremer, Jerome de Seze, François Dejardin, Stéphane Petres, Rhea Longley, Marija Backovic, Ivo Mueller, Michael T White

## Abstract

**Background:** Infection with SARS-CoV-2 induces an antibody response targeting multiple antigens that changes over time. This complexity presents challenges and opportunities for serological diagnostics.

**Methods:** A multiplex serological assay was developed to measure IgG and IgM antibody responses to seven SARS-CoV-2 spike or nucleoprotein antigens, two antigens for the nucleoproteins of the 229E and NL63 seasonal coronaviruses, and three non-coronavirus antigens. Antibodies were measured in serum samples from patients in French hospitals with RT-qPCR confirmed SARS-CoV-2 infection (*n* = 259), and negative control serum samples collected before the start of the SARS-CoV-2 epidemic (*n* = 335). A random forests algorithm was trained with the multiplex data to classify individuals with previous SARS-CoV-2 infection. A mathematical model of antibody kinetics informed by prior information from other coronaviruses was used to estimate time-varying antibody responses and assess the potential sensitivity and classification performance of serological diagnostics during the first year following symptom onset. A statistical estimator is presented that can provide estimates of seroprevalence in very low transmission settings.

**Results:** IgG antibody responses to trimeric Spike protein identified individuals with previous RT-qPCR confirmed SARS-CoV-2 infection with 91.6% sensitivity (95% confidence interval (CI); 87.5%, 94.5%) and 99.1% specificity (95% CI; 97.4%, 99.7%). Using a serological signature of IgG and IgM to multiple antigens, it was possible to identify infected individuals with 98.8% sensitivity (95% CI; 96.5%, 99.6%) and 99.3% specificity (95% CI; 97.6%, 99.8%). Informed by prior data from other coronaviruses, we estimate that one year following infection a monoplex assay with optimal anti-S^tri^ IgG cutoff has 88.7% sensitivity (95% CI: 63.4%, 97.4%), and that a multiplex assay can increase sensitivity to 96.4% (95% CI: 80.9%, 100.0%). When applied to population-level serological surveys, statistical analysis of multiplex data allows estimation of seroprevalence levels less than 1%, below the false positivity rate of many other assays.

**Conclusion:** Serological signatures based on antibody responses to multiple antigens can provide accurate and robust serological classification of individuals with previous SARS-CoV-2 infection. This provides potential solutions to two pressing challenges for SARS-CoV-2 serological surveillance: classifying individuals who were infected greater than six months ago, and measuring seroprevalence in serological surveys in very low transmission settings.

## Introduction

Severe acute respiratory syndrome coronavirus 2 (SARS-CoV-2) causing coronavirus disease 2019 (COVID-19) emerged in Wuhan, China in December 2019. Since then, it has spread rapidly, with confirmed cases being recorded in nearly every country in the world. The presence of viral infection can be directly detected via reverse transcriptase quantitative PCR (RT-qPCR) on samples from nasopharyngeal or throat swabs. For individuals who display symptoms, SARS-CoV-2 virus is detectable in the first 2-3 weeks following symptom onset [1,2]. Viral shedding is shorter in mild cases with only upper respiratory tract symptoms (1-2 weeks) [3]. For asymptomatic individuals, the duration for which SARS-CoV-2 virus can be detected is uncertain. In most countries neither mild cases nor asymptomatic cases will be tested by RT-qPCR (unless they are direct contacts of known cases), and even among tested individuals many may be viremia negative at time of testing due to low viral load or improper sampling. While not suitable for diagnosis of clinical cases, serology is a promising tool for identifying individuals with previous infection by detecting antibodies generated in response to SARS-CoV-2. However, the utility of serological testing depends on the kinetics of the anti-SARS-CoV-2 antibody response during and after infection.

An individual is seropositive to a pathogen if they have detectable antibodies specific for that pathogen. From an immunological perspective, an individual can be defined as seropositive if they have either antibody secreting plasma cells and/or a matured memory B cell response to antigens on that pathogen. In practice, serological assays are used to measure antibody responses in blood samples. However, individuals who have never been infected with the target pathogen may have non-zero antibody responses due to cross-reactivity with other pathogens or background assay noise. To account for this, defining seropositivity is equivalent to determining whether the measured antibody responses is greater or lower than some defined cutoff value [4].

The most fundamental measure of antibody level is via concentration in a sample (e.g. in units of μg/mL), however a measurement in terms of molecular mass per volume is usually impossible to obtain. Instead, a range of assays can provide measurements that are positively associated with the true antibody concentration, e.g. an optical density from an enzyme-linked immunosorbent assay (ELISA), or a median fluorescent intensity (MFI) from a Luminex^®^ microsphere assay. In contrast to the continuous measurement of antibody response provided by laboratory-based research assays, most point-of-care serological tests provide a binary outcome: seronegative or seropositive. There are several commercially available tests for detecting SARS-CoV-2 antibody responses, which are being catalogued by FIND Diagnostics [5]. These tests are typically based on lateral flow assays mounted in plastic cartridges which detect antibodies in small volume blood samples. A key feature of many rapid tests is that they are dependent on the choice of seropositivity cutoff, and there may be substantial misclassification for antibody levels close to this cutoff.

Antibody levels are not constant, and change over time. The early kinetics of the antibody response to SARS-CoV-2 have been well documented with a rapid rise in antibody levels occurring 5-15 days after symptom onset leading to seroconversion (depending on the choice of cutoff) [1,6-9]. There are not yet data on the long-term kinetics of the SARS-CoV-2 antibody response. Assuming the antibody response is similar to that of other pathogens [10-14], we expect to observe a bi-phasic pattern of decay, with rapid decay in the first 3-6 months after infection, followed by a slower rate of decay. Notably, this decay pattern may lead to seroreversion whereby a previously seropositive individual reverts to being seronegative. If a serological test with an inappropriately high choice of cutoff is used for SARS-CoV-2 serological surveys, there is a major risk that seroreversion may lead to previously infected individuals testing seronegative [15].

The antibody response generated following SARS-CoV-2 infection is diverse, consisting of multiple isotypes targeting several proteins on the virus including the spike protein (and its receptor binding domain, RBD) and the nucleoprotein [16]. This complexity of biomarkers provides both a challenge and an opportunity for diagnostics research. The challenge lies in selecting appropriate biomarkers and choosing between the increasing number of commercial assays, many of which have not been extensively validated and may produce conflicting results. The opportunity is that with multiple biomarkers, it is possible to generate a serological signature of infection that is robust to how antibody levels change over time [17-20], rather than relying on classification of seropositive individuals using a single cutoff antibody level.

In this analysis, we apply mathematical models of antibody kinetics to serological data from the early stages of SARS-CoV-2 infection and predict the potential consequences for serological diagnostics within the first year following infection.

## Methods

### Samples

We analysed 97 serum samples from 53 patients admitted to hospitals in Paris with SARS-CoV-2 infection confirmed by RT-qPCR [21,22], and 162 serum samples from healthcare workers in hospitals in Strasbourg [23] (Table 1). 68 plasma samples from the Thai Red Cross, 90 serum samples from Peruvian healthy donors, and 177 serum samples from French blood donors collected before December 2019 were used as negative controls. All samples underwent a viral inactivation protocol by heating at 56 °C for 30 minutes. The potential effect of the viral inactivation protocol on the measurement of antibody levels was assessed using serum positive for anti-malaria antibodies. IgG and IgM antibody levels were measured in matched samples before and after the inactivation protocol. The viral inactivation protocol did not affect measured IgG or IgM levels (data not shown).

**Table 1:** Panels of sasmples. Positive control serum samples are from patients with RT-qPCR confirmed SARS-CoV-2 infection. Negative control samples are from panels of pre-epidemic cohorts with ethical approval for broad antibody testing. Age is presented as median and range.

| Panel | RT-qPCR confirmed | N: participants | N: samples | age (years) | symptoms |  |
| --- | --- | --- | --- | --- | --- | --- |
|  |  |  |  |  | mild | severe |
| Hôpital Bichat, Paris | yes | 4 | 34 | 39 (31, 80) | 0 | 4 |
| Hôpital Cochin, Paris | yes | 49 | 63 | 56 (26, 79) | 27 | 22 |
| Nouvel Hôpital Civil & Hôpital de Haute Pierre, Strasbourg | yes | 162 | 162 | 32 | 162 | 0 |
| Thai Red Cross | pre-epidemic negative controls | 68 | 68 | > 18 |  |  |
| Peru negative controls | pre-epidemic negative controls | 90 | 90 | > 18 |  |  |
| France blood donors (Établissement Français du Sang) | pre-epidemic negative controls | 177 | 177 | > 18 |  |  |

### Serological assays

In a first step, four proteins derived from SARS-CoV-2 Spike were included in the assay. This includes SARS-CoV-2 trimeric Spike ectodomain (S^tri^) and its receptor-binding domain (RBD) produced as recombinant proteins in mammalian cells in the Structural Virology Unit at Institut Pasteur, while S1 (cat# REC31806) and S2 (cat# REC31807) subunits were purchased from Native Antigen, Oxford, UK. S^tri^ and RBD were designed based on the viral genome sequence of the SARS-CoV-2 strain France/IDF0372/2020, obtained from the GISAID database (accession number EPI_ISL_406596). The synthetic genes, codon-optimized for protein expression in mammalian cells, were ordered from GenScript and cloned in pcDNA3.1(+) vector as follows: the RBD, residues 331-519, and the entire S ectodomain (residues 1-1208). The RBD construct included an exogenous signal peptide of a human kappa light chain (METDTLLLWVLLLWVPGSTG) to ensure efficient protein secretion into the media. The S ectodomain construct was engineered, as reported before to have the stabilizing double proline mutation (KV986-987 to PP986-987) and the foldon domain at the C-terminus that allows the S to trimerize (YIPEAPRDGQAYVRKDGEWVLLSTFL) resembling the native S state on the virion [24]. Both constructs contained a Strep (WSHPQFEK), an octa-histidine, and an Avi tag (GLNDIFEAQKIEWHE) at the C-terminus for affinity purification. Protein expression was done by transient transfection of mammalian HEK293 free style cells, as already reported. Proteins were then purified from supernatants on a Streptactin column (IBA Biosciences) followed by size exclusion purification on Superdex 200 column using standard chromatography protocols.

In a second step, eight proteins were added to the assay. Recombinant SARS-CoV-2 nucleoprotein (NP) was expressed in *E. coli* in the Production and Purification of Recombinant Proteins Technological Platform at Institut Pasteur. Two SARS-CoV-2 antigens were purchased from Native Antigen, Oxford, UK: RBD (cat# REC31831-20) and NP (cat# REC31812-100). Additional antigens for seasonal coronaviruses 229E NP (cat# REC31758-100) and NL63 NP (cat# REC31759-100), influenza A (cat# FLU-H1N1-HA-100), adenovirus type 40 (cat# NAT41552-100) and rubella (cat# REC31651-100) were purchased from Native Antigen. All proteins were coupled to magnetic beads as described elsewhere [25]. The mass of proteins coupled on beads were optimized to generate a log-linear standard curve with a pool of positive serum prepared from RT-qPCR-confirmed SARS-CoV-2 patients.

In total, we optimized a 12-plex assay able to detect antibody responses against seven SARS-CoV-2 antigens (two nucleoproteins constructs, five spike), one nucleoprotein for each seasonal coronavirus NL63 and 229E, and three antigens from other viruses (Influenza A H1N1, adenovirus type 40, rubella) for which a large part of the population is expected to be seropositive due to vaccination or natural infection and hence serve as internal controls (Supplementary Table 1).

The assay was performed in black, 96 well, non-binding microtiter plate (cat#655090; Greiner Bio-One, Germany). Briefly 50 µL of protein-conjugated magnetic beads (500/region/µL) and 50 µL of diluted serum were mixed and incubated for 30 min at room temperature on a plate shaker. All dilutions were made in phosphate buffered saline containing 1% bovine serum albumin and 0.05% (v/v) Tween-20 (denoted as PBT), and all samples were run in singlicate. Following incubation, the magnetic beads were separated using magnetic plate separator (Luminex^®^) for 60 seconds and washed three times with 100 μL of PBT. The washed magnetic beads were incubated for 15 minutes with detector secondary antibody at room temperature on a plate shaker. The magnetic beads were separated and washed three times with 100 μL of PBT and finally resuspended in 100 μL of PBT. For IgM measurements, serum samples were diluted 1/200, and R-Phycoerythrin (R-PE) - conjugated Donkey Anti-Human IgM (cat#709-116-073; JacksonImmunoResearch, UK) antibody was used as secondary antibody at 1/400 dilution. For IgG, serum samples were diluted 1/100, and R-Phycoerythrin (R-PE) -conjugated Donkey Anti-Human IgG (cat#709-116-098; JacksonImmunoResearch, UK) antibody was used as secondary antibody at 1/120 dilution.

On each plate, two blanks (only beads, no serum) were included as well as a standard curve prepared from two-fold serial dilutions (1:50 to 1:25600) of a pool of positive controls. Plates were read using a Luminex^®^ MAGPIX^®^ system and the median fluorescence intensity (MFI) was used for analysis. A 5-parameter logistic curve was used to convert MFI to antibody dilution, relative to the standard curve performed on the same plate to account for inter-assay variations. The multiplex immunoassay was validated by checking that the MFI obtained were well correlated with those obtained in monoplex (only one conjugated bead type per well). For non-SARS-CoV-2 antigens, MFI data was used for the analysis.

### Statistical evaluation of diagnostic performance

For measured antibody responses to a single antigen, diagnostic sensitivity is defined as the proportion of patients with RT-qPCR confirmed SARS-CoV-2 infection with measured antibody levels above a given seropositivity cutoff. For assessment of classification performance, samples taken from individuals less than 10 days after symptom onset were excluded. Diagnostic specificity is defined to be the proportion of negative controls (with no history of SARS-CoV-2 infection) with measured antibody levels below a given seropositivity cutoff. Sensitivity and specificity can be traded off against each other by varying the seropositivity cutoff. This trade-off is formally evaluated using Receiver Operating Characteristic (ROC) analysis.

Measured antibody responses to multiple antigens can be combined to identify individuals with previous SARS-CoV-2 infection using classification algorithms. Here we use a random forests algorithm [17]. Uncertainty in sensitivity and specificity is quantified in three ways: (i) binomial confidence intervals calculated using Wilson’s method; (ii) 1000-fold repeat cross-validation with a training set comprising 2/3 of the data and a disjoint testing set comprising 1/3 of the data; (iii) cross-panel validation with algorithms trained and tested on disjoint panels of data (Supplementary Figure S1).

### Mathematical model of antibody kinetics

SARS-CoV-2 antibody kinetics are described using a previously published mathematical model of the immunological processes underlying the generation and waning of antibody responses following infection or vaccination [10]. The existing model is adapted to account for the frequent data available in the first weeks of infection.

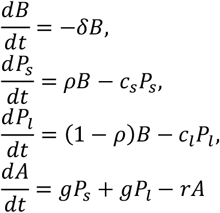

where *B* denotes B lymphocytes, δ is the rate of differentiation of B lymphocytes into antibody secreting plasma cells, *P*_*s*_ denotes short-lived plasma cells, *P*_*l*_ denotes long-lived plasma cells, *ρ* is the proportion of plasma cells that are short-lived, *g* is the rate of generation of antibodies (IgG or IgM) from plasma cells, and *r* is the rate of decay of antibody molecules. Assuming *B*(0) = *B*_*0*_, *P*_*s*_(0) = *P*_*l*_(0) = 0 and *A*(0) = *A*_*bg*_, these equations can be solved analytically to give:

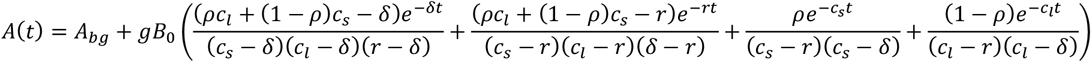

Statistical inference was implemented within a mixed-effects framework allowing for characterisation of the kinetics within each individual while also describing the population-level patterns. On the population level, both the mean and variation in antibody kinetics are accounted for. The models were fitted in a Bayesian framework using Markov chain Monte Carlo methods with informative priors. Posterior parameter estimates are presented as medians with 95% credible intervals (CrIs).

### Prior data

The recent emergence of SARS-CoV-2 means that long-term data on the duration of antibody responses do not yet exist. Therefore, predictions of antibody levels beyond the period for which data has been collected are heavily dependent on structural model assumptions and assumed prior information. The prior estimate of the half-life of IgG molecules is 21 days. The prior estimate of the half-life of IgM molecules is 10 days. Prior estimates for the short-lived component of the antibody response (half-life = 3.5 days) are consistent with data from several sources [10-14]. The most notable uncertainty relates to estimates of the duration of the long-lived component of the SARS-CoV-2 antibody responses. We reviewed data from a number of sources on the long-term antibody kinetics following infection with other coronaviruses [26-31], summarized in Appendix Table A1. Based on the wide range of long-term antibody kinetics observed, we assumed a prior estimate of the half-life of the long-lived component of the IgG antibody response to be 400 days, and that the proportion of the short-lived antibody secreting cells is 90%. This corresponds to a scenario where the IgG antibody responses decreases by approximately 60% after one year. Additional sensitivity analyses were run assuming the half-life of the long-lived component of the IgG antibody response to be 200 days and 800 days.

The model was first fitted to data from 23 patients with RT-qPCR confirmed SARS-CoV-2 infection in Hong Kong hospitals who were followed longitudinally for up to four weeks after initial onset of symptoms [1]. Posterior estimates from this model and data were used to provide prior estimates for the parameters describing the early stages of the antibody response (Appendix Table A2).

### Serological surveillance

A ROC curve obtained from a training data set consisting of both positive and negative samples is described by a sequence of sensitivities and specificities 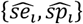 N-fold cross-validation generates samples of sensitivity {*se*_*i*1_, …, *se*_*iN*_} for each 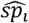 and samples of specificity {*sp*_*i*1_, …, *sp*_*iN*_} for each 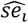 Following a previously outlined approach [32,33], for each pair *i* of sensitivity and specificity, we obtain *N* estimates of the measured seroprevalence *M*_*in*_ in a scenario with true seroprevalence *T* as follows:

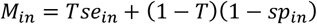

The point estimates of sensitivity and specificity can be used to calculate an adjusted estimate of true seroprevalence:

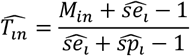

With 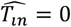 if 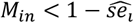 Both *M*_*i*_ and 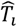 are summarized as medians with 95% ranges. We calculate the expected relative error as:

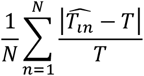

### Ethics

Serum samples were obtained through the CORSER study (Etude séro-épidémiologique du virus SARS-CoV-2 en France : constitution d’une collection d’échantillons biologiques humains) directed by Institut Pasteur and approved by the Comité de Protection des Personnes Ile de France III, and the French COVID cohort (NCT04262921, sponsored by Inserm and approved by the Comité de Protection des Personnes Ile de France VI). Sample collection in Hôpital Cochin was approved by the Research Ethics Commission of Necker-Cochin Hospital. Use of the Peruvian negative controls was approved by the Institutional Ethics Committee from the Universidad Peruana Cayetano Heredia (SIDISI 100873).

## Results

### Single biomarker classification

IgG and IgM antibody responses to twelve antigens were measured as median fluorescence intensity (MFI). For the seven SARS-CoV-2 antigens, the measured MFI was converted to antibody dilutions (Figure 1). For all 14 SARS-CoV-2 biomarkers (seven antigens, IgG and IgM for both), measured responses were significantly higher in samples with RT-qPCR confirmed infection than in negative control samples (Figure 1A-B; P value < 1 × 10^−7^; 2 sided t test).

**Figure 1:**
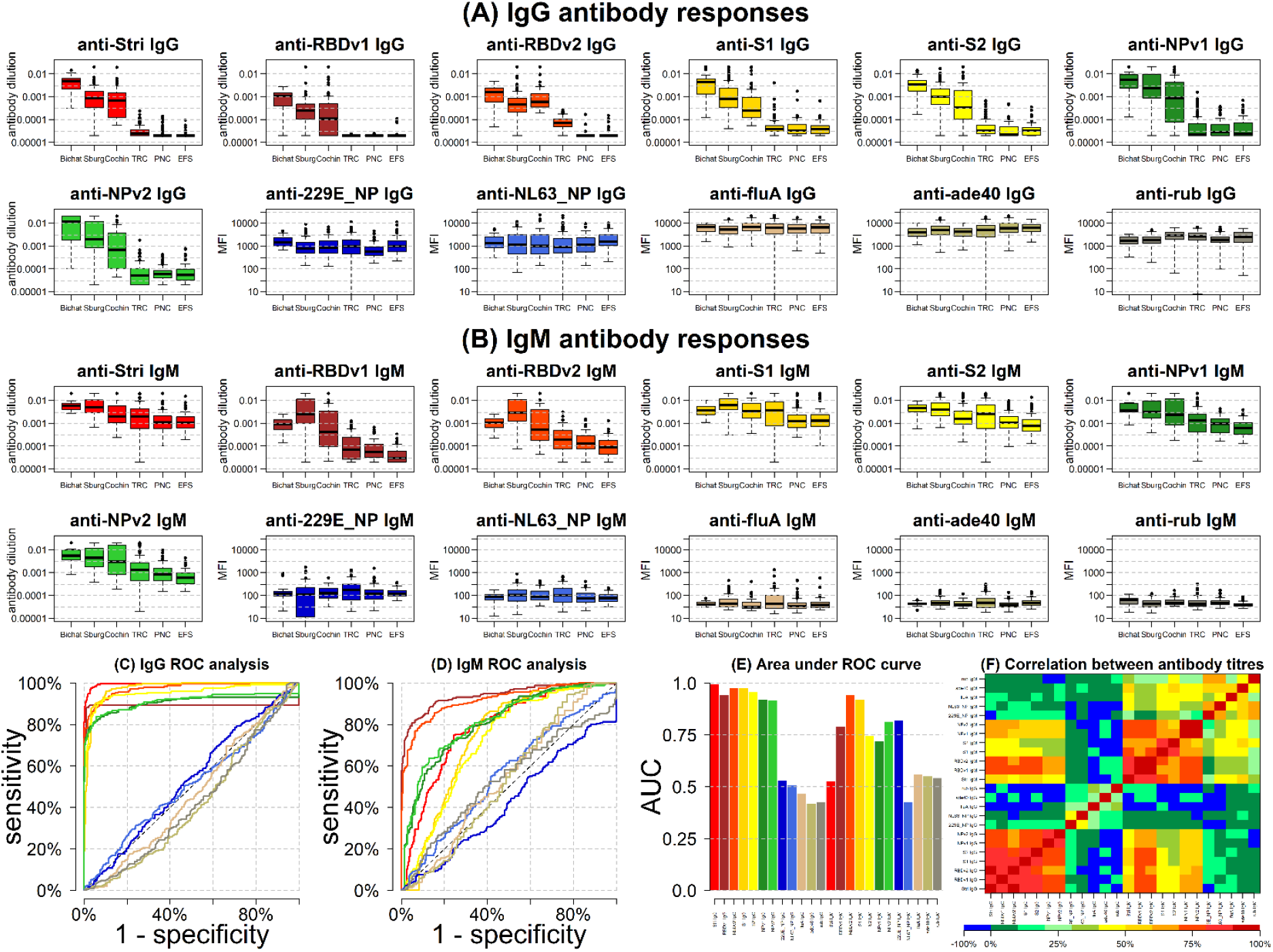
Anti-SARS-CoV-2 antibody responses. **(A)** Measured IgG antibody dilutions or medium fluorescence intensity (MFI) in serum samples with previously confirmed RT-qPCR infection from patients in Hôpital Bichat (*n* = 34), health care workers from Strasbourg (*n* = 162), and Hôpital Cochin (*n* = 63). Negative control samples from Thailand (*n* = 68), Peru (*n* = 90), and French donors (*n* = 177) were also tested. **(B)** Measured IgM antibody dilutions or MFI in serum or plasma samples. **(C)** Receiver Operating Characteristic (ROC) curve for IgG antibodies obtained by varying the cutoff for seropositivity. Colours correspond to those shown in part A. **(D)** ROC curve for IgM antibodies obtained by varying the cutoff for seropositivity. **(E)** Area under the ROC curve for individual biomarkers. **(F)** Spearman correlation between measured antibody responses.

The trade-off between sensitivity and specificity obtained by varying the cutoff for seropositivity was investigated using a ROC curve (Figure 1C-D). Depending on the characteristics of the desired diagnostic test, different targets for sensitivity and specificity can be considered. The results of three targets are summarized in Table 2. These are: (i) high sensitivity target enforcing sensitivity > 99%; (ii) balanced sensitivity and specificity where both are approximately equal; and (iii) high specificity target enforcing specificity >99%. Focusing on the high specificity target, anti-S^tri^ IgG was the best performing biomarker with 99.1% specificity (95% CI: 97.4%, 99.7%) and 91.6% sensitivity (95% CI: 87.5%, 94.5%). Anti-S^tri^ IgG provided significantly better classification than all other biomarkers (Supplementary Table S2; McNemar’s test P value < 10^−8^). There was significant correlation between antibody responses against all SARS-CoV-2 antigens, but no significant correlation between antibody responses to SARS-CoV-2 and the seasonal coronaviruses 229E and NL63 (Figure 1E).

**Table 2:**
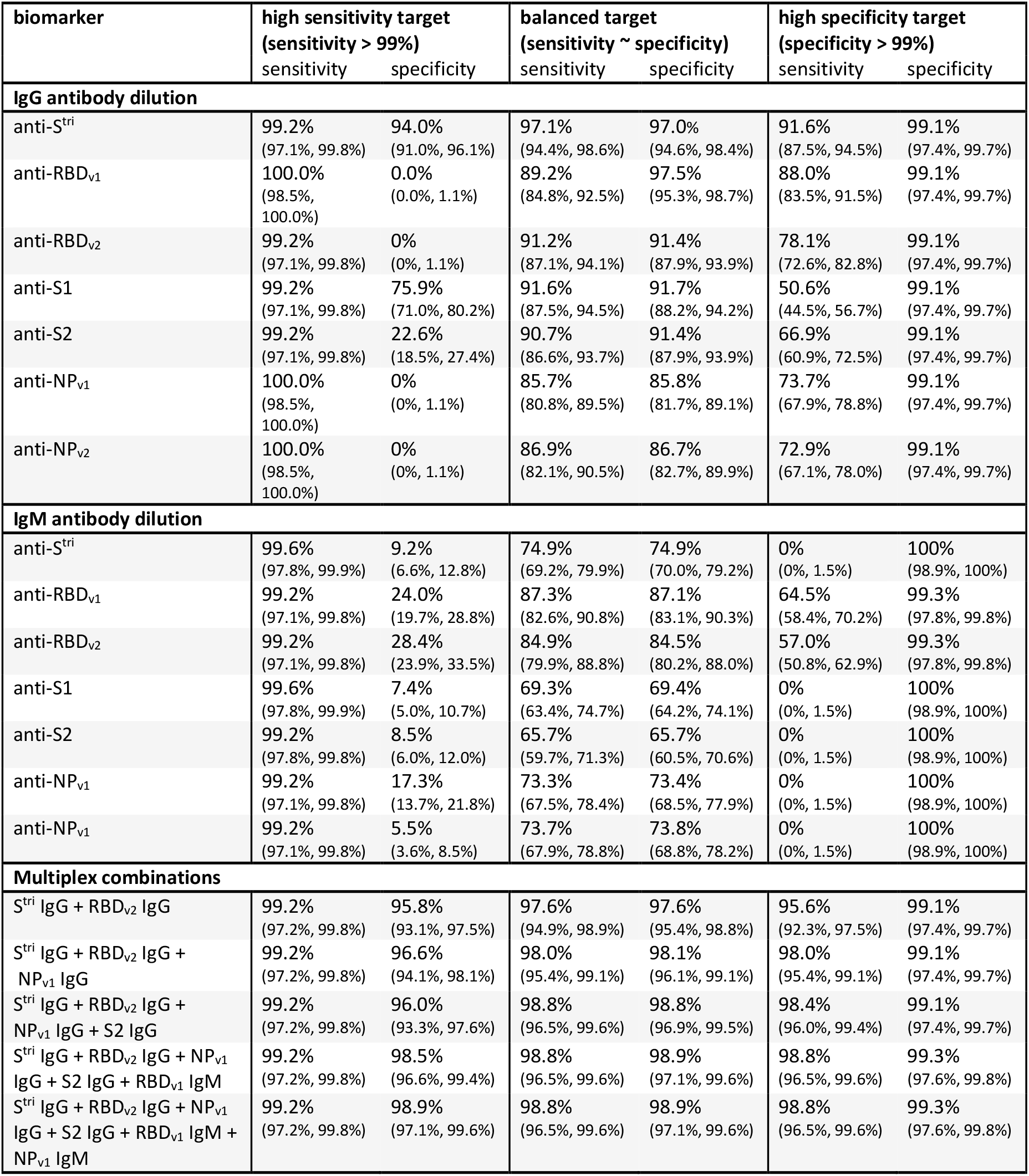
Sensitivity and specificity targets for single biomarkers and multiplex combinations. 95% binomial confidence intervals were calculated using Wilson’s method. Antigen combinations were selected to optimize sensitivity for the high specificity target, i.e. the highest sensitivity while enforcing specificity > 99%.

### Serological signatures and multiple biomarker classification

With 24 biomarkers, there are 24*23/2 = 156 possible pairwise comparisons. Figure 2A provides an overview of six pairwise comparisons of antibody responses. The data are noisy, highly correlated and high dimensional (although only two dimensions are depicted here). We refer to the pattern of multiple antibody responses in multiple dimensions as the serological signature. For all plots of SARS-CoV-2 biomarkers there are two distinct clusters: antibody responses from negative control samples in blue cluster in the bottom left, and antibody responses from serum samples from individuals with RT-qPCR confirmed SARS-CoV-2 infection cluster in the centre and top right.

**Figure 2:**
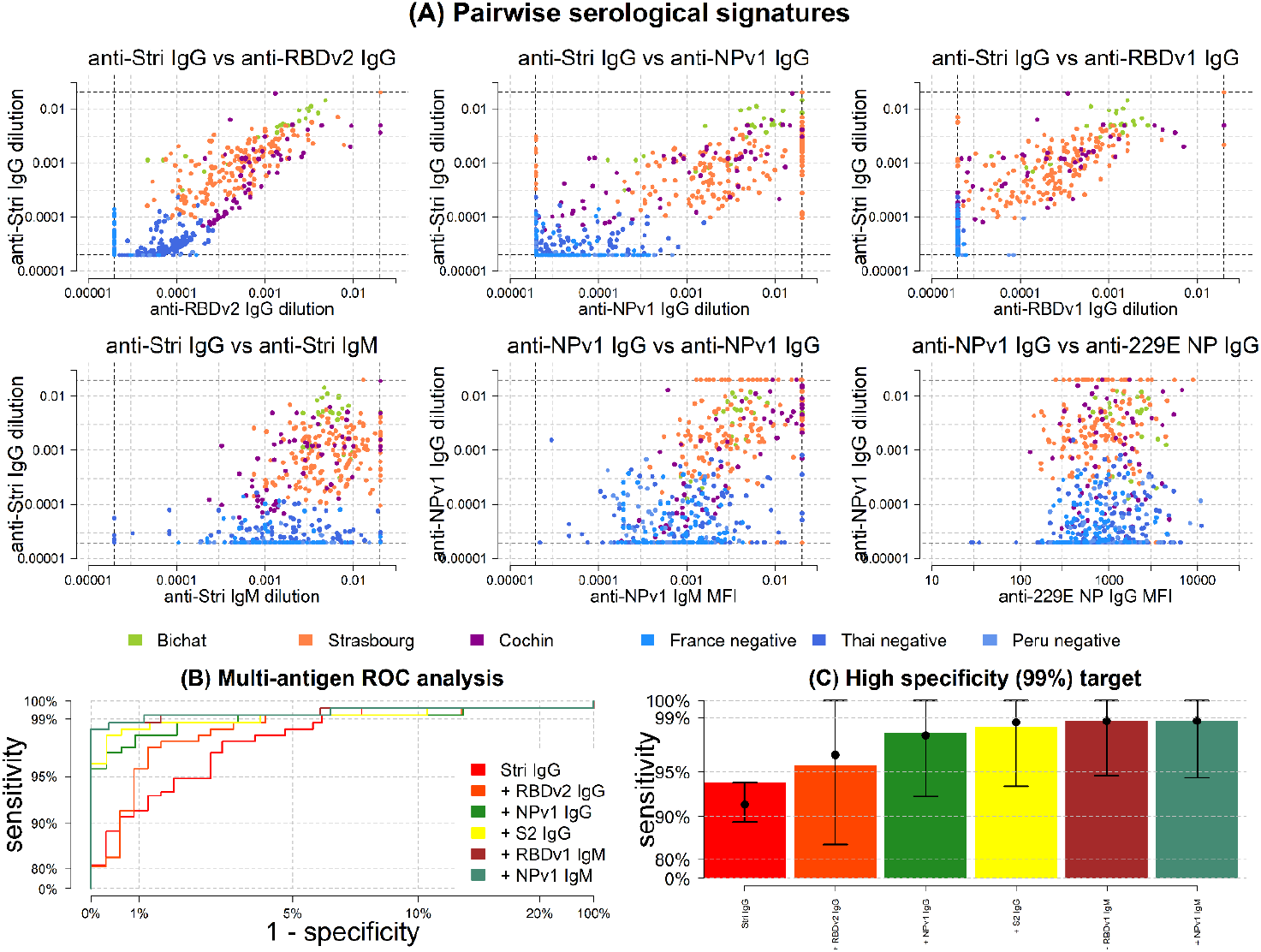
Serological signatures of SARS-CoV-2 infection. **(A)** Pairwise combinations of antibody responses. Each point denotes a measured antibody response from a sample from Hôpital Bichat (*n* = 34), Nouvel Hôpital Civil & Hôpital de Haute Pierre in Strasbourg (*n* = 162), and Hôpital Cochin (*n* = 63). Negative control samples are included from Thailand (*n* = 68), Peru (*n* = 90) and French blood donors (*n* =177). **(B)** ROC curves for multiple biomarker classifiers generated using a Random Forests algorithm. **(C)** For a high specificity target (>99%), sensitivity increases with additional biomarkers. Sensitivity was estimated using a Random Forests classifier. Points and whiskers denote the median and 95% confidence intervals from repeat cross-validation.

The classification performance of multiplex combinations of antibody responses is shown with the ROC curves in Figure 2B. Including data from additional biomarkers leads to significant improvements in classification performance (Table 2). For example, for the high specificity target, with a single biomarker (anti-S^tri^ IgG) we can achieve 91.6% sensitivity (95% CI: 87.5%, 94.5%). Including anti-RBDv2 IgG increases sensitivity to 95.6% sensitivity (95% CI: 92.3%, 97.5%). Combinations of size five to six provide 98.8% sensitivity (95% CI: 96.5%, 99.6%) and 99.3% sensitivity (95% CI: 97.6%, 99.8%). There are diminishing returns to increasing the number of additional antigens (Figure 2C).

### SARS-CoV-2 antibody kinetics

A mathematical model of antibody kinetics was fit to the serological data. Figure 3A shows data from a patient from Hôpital Bichat with frequent longitudinal sampling. The data and model indicate that the antibody response is in a rising phase between 5 and 30 days after symptom onset. The seroconversion time depends on the seropositivity cutoff. For the cutoffs shown, seroconversion occurs for anti-S^tri^ IgG, anti-RBD_v1_ IgG and anti-S2 IgG, but not for anti-RBD_v2_ IgG, anti-S1 IgG, anti-NP_v1_ IgG and anti-NP_v2_ IgG.

**Figure 3:**
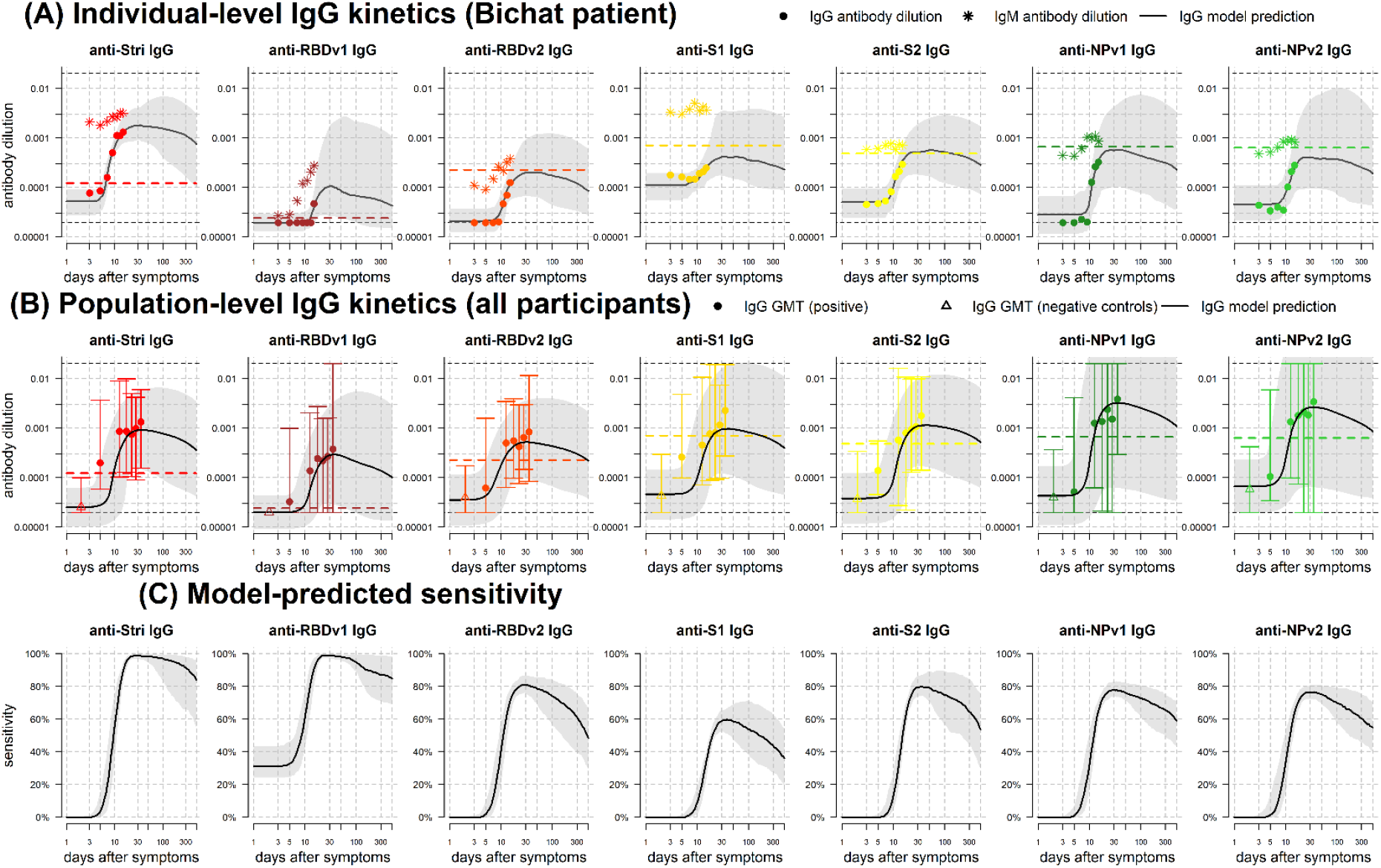
IgG antibody kinetics. **(A)** Measured IgG antibody dilutions, shown as points, from a patient in Hôpital Bichat followed longitudinally. Posterior median model predictions of IgG antibody dilution are shown as black lines, with 95% credible intervals in grey. The coloured dashed line represents the cutoff for IgG seropositivity for that antigen. IgM antibody dilutions are shown as asterisks. The black horizontal dashed lines represent the upper and lower limits of the assay. **(B)** Measured IgG antibody dilutions and model predictions for the full population. Measured IgG antibody dilutions are shown as geometric mean titre (GMT) with 95% ranges. **(C)** Model predicted proportion of individuals testing seropositive over time.

For all 215 individuals with RT-qPCR SARS-CoV-2 infection, Figure 3B shows the model predicted IgG antibody response to SARS-CoV-2. For all antigens, we predict a bi-phasic pattern of waning with a first rapid phase between one and three months after symptom onset, followed by a slower rate of waning. The percentage reduction in antibody level after one year was mostly determined by prior information and estimated to be 47% (95% CrI: 18%, 90%) for anti-S^tri^ IgG antibodies, with comparable estimates for other antigens (Appendix Table A3).

Sensitivity was assessed using the seropositivity cutoff based on the high specificity target in Table 2. For all antigens considered, we predict that there will be a reduction in sensitivity over time, although there is a large degree of uncertainty (Figure 3C). In particular, we predict that the sensitivity based on anti-S^tri^ IgG antibody responses after one year will be 88.7% (95% CrI: 63.4%, 97.4%); and that the sensitivity of a four antigen multiplex classifier after six months will be 96.4% (95% CrI: 80.9%, 100%) (Figure 4).

**Figure 4:**
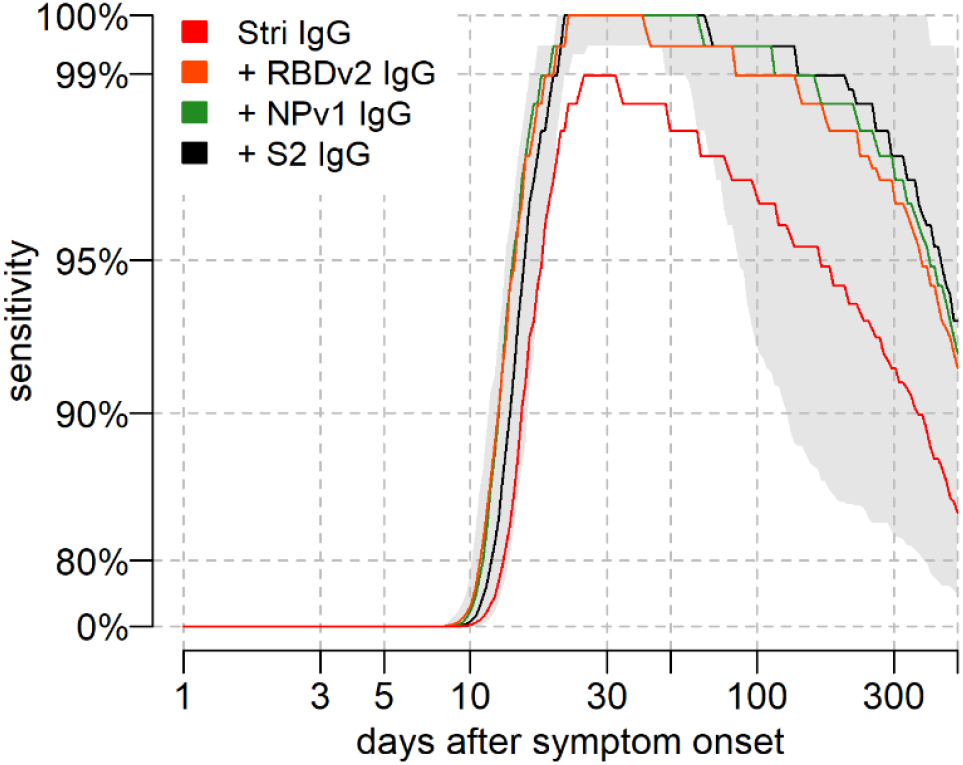
Model predicted sensitivity over time. Proportion of *n* = 215 individuals with qRT-PCR infection testing seropositive over time. A Random Forests algorithm was used for classification of multiple antigen multiplex data. The grey shaded region shows the 95% uncertainty interval for the four antigen multiplex classifier.

### Multiplex assays for seroprevalence surveys

For serological diagnosis of individual samples, the target pursued thus far is to optimize sensitivity whilst enforcing high specificity (>99%). A serological assay that accurately classifies individual samples will also perform well at estimating seroprevalence in populations. However, an assay optimized for individual-level classification is not necessarily optimal for population-level surveillance where the target is to obtain accurate estimates of the true seroprevalence. Figure 5A presents ROC curves for a monoplex anti-S^tri^ IgG assay and a multiplex assay using six biomarkers from Table 2 with quantification of uncertainty via repeat cross-validation. In an epidemiological scenario with true seroprevalence = 5%, the measured seroprevalence will depend on the assay sensitivity and false positive rate (= 1 – specificity) (Figure 5B). For high false positive rate, the measured seroprevalence overestimates the true seroprevalence. Applying a statistical correction to account for imperfect sensitivity and specificity, we can obtain more accurate estimates of seroprevalence (Figure 5C). For both the monoplex and multiplex serological assays, the adjusted estimates are not accurate for high false positive rates.

**Figure 5:**
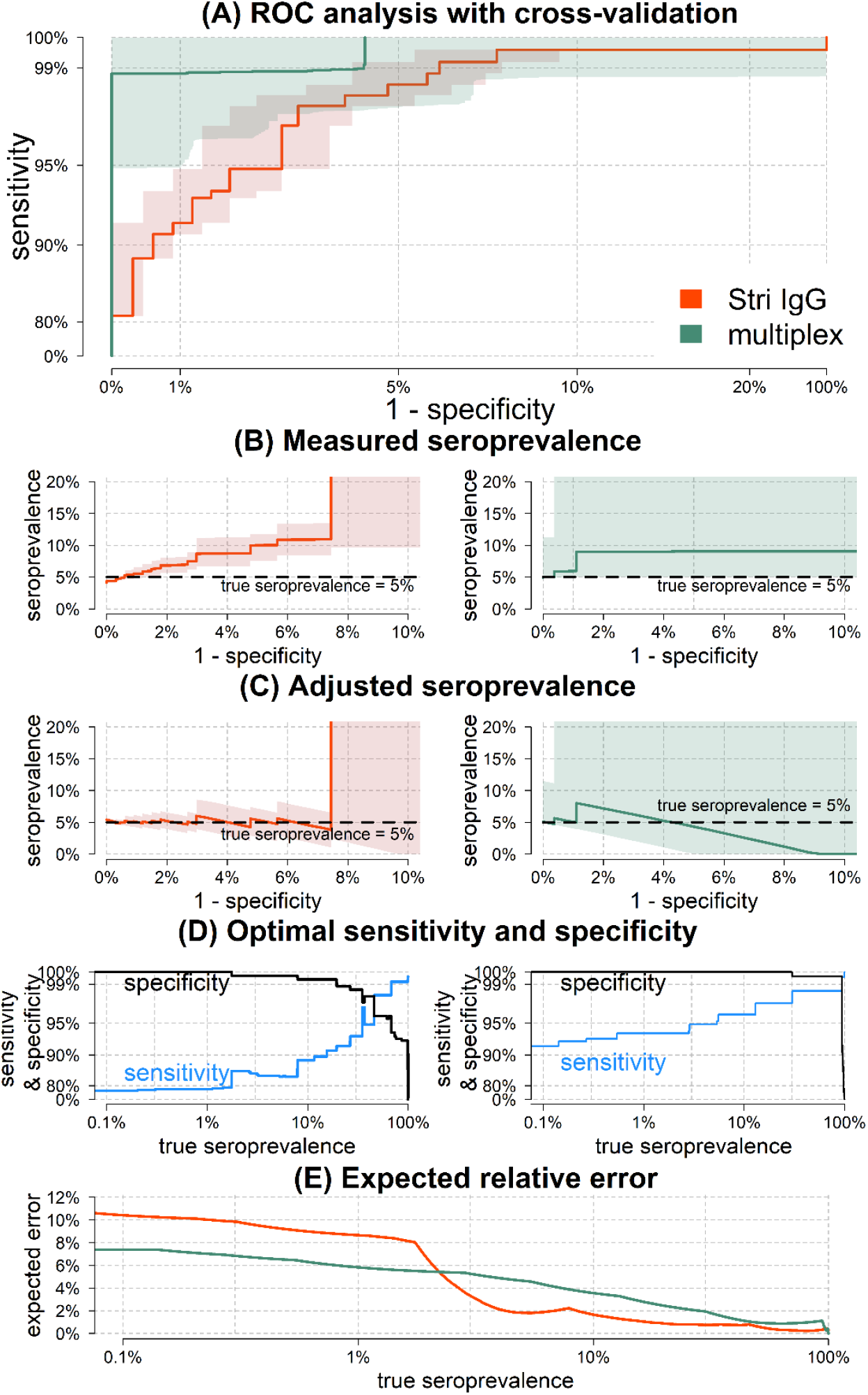
Implementation of seroprevalence surveys. **(A)** Receiver Operating Characteristic (ROC) analysis with cross-validated uncertainty. Solid lines represent median ROC curves and shaded regions represent 95% uncertainty intervals for specificity. **(B)** In a scenario with true seroprevalence = 5%, the measured seroprevalence depends on the false positive rate (= 1 – specificity). Results for the monoplex anti-S^tri^ IgG assay are shown on the left, and results for the multiplex assay are shown on the right. **(C)** In a scenario with true seroprevalence = 5%, adjusted seroprevalence estimates are obtained by accounting for assay sensitivity and specificity. **(D)** Across a range of true seroprevalence, optimal values of sensitivity and specificity can be selected to minimize the expected relative error in seroprevalence surveys. **(E)** The expected relative error for optimal values of sensitivity and specificity.

Figure 5B-C presents the scenario when seroprevalence is known to be 5%. In real applications, true seroprevalence is not known *a priori*. For a range of seroprevalence from 0.1% to 100%, Figure 5D presents values of the assay’s sensitivity and specificity that have been optimized to minimize the expected relative error. For a monoplex assay based on anti-S^tri^ IgG antibodies, if true seroprevalence <20% the relative error is minimized when we select specificity >99%. When true seroprevalence <2% the relative error is minimized when specificity = 100%. For a multiplex serological assay, if true seroprevalence <30%, the relative error is minimized when we implement an algorithm with specificity = 100%. Figure 5E presents a comparison of the expected relative error for the monoplex and multiplex assays. The expected relative error depends on the possible values of sensitivity and specificity, as well as the uncertainty in these estimates. For true seroprevalence >2% the monoplex assay has lower error (a consequence of the lower levels of variation in the ROC curve). For true seroprevalence <2%, the multiplex assay has lower error, a consequence of the high levels of specificity.

## Discussion

Infection with SARS-CoV-2 induces antibodies of multiple isotypes (IgG, IgM, IgA) targeting multiple epitopes on spike proteins exposed on the virus surface, and nucleoprotein. Each of these biomarkers may exhibit distinct kinetics leading to variation in their potential diagnostic performance. There is also substantial between-individual variation in the antibody response generated following SARS-CoV-2 infection. By measuring multiple biomarkers in large numbers of individuals, it is possible to create a serological signature of previous infection [17-19]. Although necessarily more complex than a single measured antibody response, such an approach has the potential of providing more accurate classification and being more stable over time.

IgG antibody levels to a single antigen (trimeric Spike) can classify samples from individuals previously infected with SARS-CoV-2 with 91.6% sensitivity (95% CI: 87.5%, 94.5%) and 99.1% specificity (95% CI: 97.4%, 99.7%). Measuring additional biomarkers with a multiplex assay can improve classification performance to 98.8% sensitivity (95% CI: 96.5%, 99.6%) and 99.3% specificity (95% CI: 97.6%, 99.8%). A similar phenomenon is observed for serological diagnosis of HIV where combining multiple assays can lead to improved accuracy [34]. Multiplex assays provide some of the benefits of combining separate assays, but are subject to the risk that multiple biomarkers measured on the same assay are often correlated. An additional role for high accuracy multiplex assays is as a secondary assay after initial screening with point-of-care rapid serological tests.

The reported accuracy of serological tests depends on multiple factors, most notably the validation samples used. Specificity is typically determined by pre-epidemic negative control samples, with the inclusion of greater numbers of samples providing more robust characterization of specificity. Rather than taking large numbers of samples from a homogeneous population, we encourage the utilization of multiple negative control panels that are epidemiologically diverse with respect to age and location. Sensitivity is determined by positive control samples. It may be trivial to record high sensitivity when validating with samples from small numbers of individuals with severe symptoms [35]. We encourage the use of multiple panels of positive control panels that are epidemiologically diverse with respect to factors such as age, COVID-19 symptom severity, and time since symptoms. When comparing the performance of different assays, the ideal approach is to use common serum samples. In the majority of situations where common serum samples are not available, including epidemiological information on validation samples can facilitate more effective comparison between assays.

The long-term kinetics of the antibody response to SARS-CoV-2 will not be definitively quantified until infected individuals are followed longitudinally for months and even years after RT-qPCR confirmed infection. As we wait for this data to be collected, mathematical models can provide important insights into how SARS-CoV-2 antibody levels may change over time. Modelling beyond the timeframe for which we have data has its limitations, however our approach benefits from robust quantification of uncertainty accounting for a wide range of future scenarios. Furthermore, this modelling approach provides falsifiable predictions which will allow models to be updated as our team and others generate new data. For the purpose of evaluation of antibody kinetics, measured antibody responses from samples collected from individuals followed longitudinally after confirmed SARS-CoV-2 infection will be especially valuable.

The simulations presented here predict that following SARS-CoV-2 infection, antibody responses will increase rapidly 1-2 weeks after symptom onset, with antibody responses peaking within 2-4 weeks. After this peak, antibody responses are predicted to decline according to a bi-phasic pattern, with rapid decay in the first three to six months followed by a slower rate of decay. Model predictions of the rise and peak of antibody response are informed by, and are consistent with, many sources of data [10-14,36]. Model predictions of the decay of antibody responses are strongly determined by prior information on longitudinal follow-up of individuals infected with other coronaviruses [26-31]. Under the scenario that the decay of SARS-CoV-2 antibody responses is similar to that of SARS-CoV, we would expect substantial reductions in antibody levels within the first year after infection. For the seropositivity cutoffs highlighted here, this could cause approximately 50% - 90% of individuals to test seronegative after one year, depending on the exact choice of biomarker and seropositivity cut-off.

This presents a potential problem for SARS-CoV-2 serological diagnostics. Most commercially available diagnostic tests compare antibody responses to a fixed seropositivity cutoff. Where these cutoffs have been validated, it is typically by comparison of serum from negative control samples collected pre-epidemic with serum from hospitalized patients in the first weeks of infection (i.e. when antibody responses are likely to be at their highest) [37,38]. If we fail to account for antibody kinetics, we risk incorrectly classifying individuals with old infections (e.g. >6 months) as seronegative. This is particularly important for point-of-care rapid serological tests with fixed cutoffs, limited dynamic range and visual evaluation. If inappropriate tests are used in seroprevalence surveys, there is a risk of substantial under-estimation of the proportion of previously infected individuals.

An advantage of continuous multiplex data is that different algorithms can be applied to the same data for different epidemiological applications. Table 2 assesses classification performance against three targets. We selected multiplex combination of antigens to optimize classification of individual samples against a target of maximizing sensitivity given a minimum specificity of 99%. However, a test that is optimal for individual-level classification is not necessarily optimal for population-level use. A recommended target for serological assays for sero-surveillance surveys is to minimize the expected error in estimated seroprevalence. For scenarios where we expect low true seroprevalence (<10%) we show that assays with high specificity (>99%) are optimal (Figure 5). Notably this provides a potential solution to the challenge of implementing sero-surveillance studies in regions where seroprevalence is expected to be lower than commonly reported false positive rates [39]. This is possible because our assay allows 100% specificity to be achieved with an accompanying reduction in sensitivity that can be statistically accounted for. In low seroprevalence settings there are additional challenges in collecting sufficient numbers of samples to ensure statistically robust estimates [40].

There are a large number of immunological assays capable of measuring the antibody response to SARS-CoV-2 including neutralization assays, ELISA, Luminex, Luciferase Immunoprecipitation System (LIPS), peptide microarrays and more [40,41]. From the perspective of quantifying protective immunity and vaccine development, functional approaches such as neutralization assays are clearly preferable. However, from a surveillance and diagnostics perspective, assays should be assessed in terms of their performance at classifying individuals with previous RT-qPCR confirmed infection. If the target is to diagnose someone, it does not matter what a biomarker does, only that it can be reliably detected in previously infected individuals and not in uninfected individuals.

Beyond diagnostics, assessment of antibody kinetics may contribute to better understanding of the immune responses generated by SARS-CoV-2 vaccines [42]. Statistical models can be used to identify immunological correlates of protection, at least according to conditions such as the Prentice criterion [43,44]. An estimated correlate of protection may take the form of a dose-response relationship, with higher antibody levels associated with greater vaccine efficacy. Under the assumption that a correlate of protection can be identified, models of antibody kinetics can be used to provide preliminary estimates of the duration of protection following vaccination or natural infection [13,45].

The analysis presented here is based on limited data, and the predictions may subsequently be contradicted as more data become available. However, the concepts outlined here of serological signatures of SARS-CoV-2 infection generated by multiplex assays, and mathematical models of antibody kinetics, allow us to plan in advance for some of the future challenges that we may face in SARS-CoV-2 serological surveillance.

## Data Availability

All data and code used for reproducing the results is freely available on GitHub: https://github.com/MWhite-InstitutPasteur/SARS_CoV_2_SeroDX_phase2.

## Author contributions

JR and SP optimized protocols and processed samples. RJL contributed to protocol development. CC and CD processed samples. SM implemented inactivation protocols. NN analysed data. MB and AM designed and produced antigens. SK, BT, SFK, and JdS collected samples. IM and MW developed the concept. MW analysed the data and wrote the manuscript.

## Acknowledgements

The French COVID cohort is supported by the REACTing consortium and by the French Directorate General for Health. Darragh Duffy, Jérôme Hadjadj and Laura Barnabei are thanked for their work on the Hôpital Cochin study. Arnaud Fontanet is thanked for critical reading of the manuscript. Dionicia Gamboa is thanked for sharing negative control samples from Peru. Jetsumon Sattabongkot is thanked for sharing negative control samples from Thailand. Marie-Noelle Ungeheuer and Blanca Liliana Perlaza are thanked for processing samples at the ICAReB platform in Institut Pasteur. Shane Mansfield and Richard Davison are thanked for their help in the solution of differential equations. We thank all patients and health care workers who kindly agreed for samples to be used for medical research purposes.

---

**Supplementary Table S1:** List of antigens included in the multiplex serological assay.

| category | short name | recombinant antigen | expression system | supplier, catalog number |
| --- | --- | --- | --- | --- |
| SARS-CoV-2 | S<sup>tri</sup> | SARS-CoV-2 Trimeric Spike protein | HEK293 | Institut Pasteur, Paris |
| SARS-CoV-2 | RBD<sub>v1</sub> | SARS-CoV-2 Spike Glycoprotein (S1) RBD | CHO | Native Antigen, REC31831-100 |
| SARS-CoV-2 | RBD<sub>v2</sub> | SARS-CoV-2 Spike Glycoprotein (S1) RBD | HEK293 | Institut Pasteur, Paris |
| SARS-CoV-2 | S1 | SARS-CoV-2 Spike Glycoprotein (S1) | HEK293 | Native Antigen, REC31806-100 |
| SARS-CoV-2 | S2 | SARS-CoV-2 Spike Glycoprotein (S2) | HEK293 | Native Antigen, REC31807-100 |
| SARS-CoV-2 | NP<sub>v1</sub> | SARS-CoV-2 Nucleoprotein | *E. coli* | Institut Pasteur, Paris |
| SARS-CoV-2 | NP<sub>v2</sub> | SARS-CoV-2 Nucleoprotein | *E. coli* | Native Antigen, REC31812-100 |
| seasonal coronavirus | 229E-NP | Human Coronavirus 229E Nucleoprotein | *E. coli* | Native Antigen, REC31758-100 |
| seasonal coronavirus | NL63-NP | Human Coronavirus NL63 Nucleoprotein | *E. coli* | Native Antigen, REC31759-100 |
| internal controls | FluA | Influenza virus H1N1 haemagglutinin recombinant antigen | HEK293 | Native Antigen, FLU-H1N1-HA-100 |
| internal controls | Ade40 | Adenovirus type 40 Hexon (capsid) | HEK293 | Native Antigen, NAT41552-100 |
| internal controls | Rub | Rubella virus-like particles (spike glycoprotein E1, spike glycoprotein E2 and Capsid protein) | HEK293 | Native Antigen, REC31651-100 |

**Supplementary Table S2:**
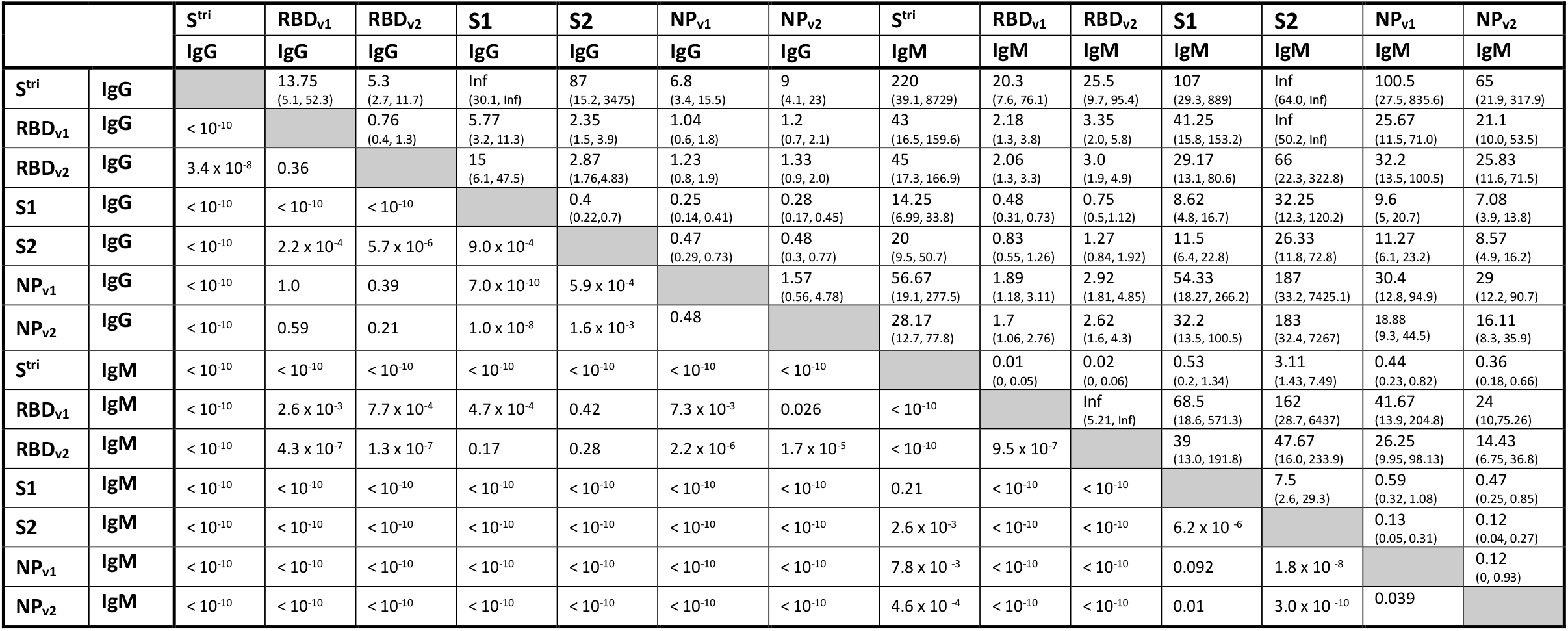
Comparison of classification performance between biomarkers for a high specificity target (>99%). Pairwise comparisons are made using McNemar’s test. The above diagonal element shows the odds ratio with 95% confidence intervals. Odds ratio > 1 indicates that biomarker indicated by the row has better classification than the biomarker indicated by the column. The corresponding element below the diagonal presents the P value.

**Supplementary Table S3:** Model predicted reductions in antibody levels and sensitivity at 6 and 12 months after symptom onset. All results are shown for a high specificity target (>99%).

|  | model predicted reduction in antibody levels |  | cutoff antibody dilution | model predicted sensitivity |  |
| --- | --- | --- | --- | --- | --- |
|  | 6 months | 12 months |  | 6 months | 12 months |
| *Single biomarkers* |  |  |  |  |  |
| S<sup>tri</sup> IgG | 23.3%<br>(3.6%, 85.4%) | 47.1%<br>(17.5%, 90.3%) | 0.000122 | 94.8%<br>(71.4%, 99.2%) | 88.7%<br>(63.4%, 97.4%) |
| RBD<sub>v1</sub> IgG | 22.1%<br>(1.9%, 94.8%) | 44.3%<br>(13.9%, 96.0%) | 0.000024 | 89.7%<br>(74.2%, 99.2%) | 86.9%<br>(70.1%, 98.7%) |
| RBD<sub>v2</sub> IgG | 23.2%<br>(3.4%, 80.4%) | 44.6%<br>(16.1%, 85.8%) | 0.000226 | 67.8%<br>(46.6%, 80.7%) | 56.7%<br>(37.9%, 73.2%) |
| S1 IgG | 20.5%<br>(2.6%, 89.9%) | 44.0%<br>(14.6%, 92.4%) | 0.000700 | 49.2%<br>(31.7%, 67.6%) | 41.2%<br>(25.5%, 59.8%) |
| S2 IgG | 18.3%<br>(2.1%, 79.5%) | 41.5%<br>(13.5%, 86.5%) | 0.000479 | 70.9%<br>(51.0%, 87.7%) | 61.6%<br>(39.9%, 81.0%) |
| NP<sub>v1</sub> IgG | 27.4%<br>(2.5%, 94.4%) | 49.4%<br>(11.5%, 96.4%) | 0.000664 | 69.0%<br>(53.6%, 78.4%) | 63.9%<br>(47.7%, 73.2%) |
| NP<sub>v2</sub> IgG | 25.5%<br>(1.6%, 97.2%) | 47.5%<br>(7.8%, 98.4%) | 0.000630 | 65.5%<br>(49.0%, 77.8%) | 57.7%<br>(43.8%, 74.3%) |
| *Multiplex combinations* |  |  |  |  |  |
| S<sup>tri</sup> IgG + RBD<sub>v2</sub> IgG | — | — | — | 97.9%<br>(89.5%, 100.0%) | 95.4%<br>(82.3%, 99.6%) |
| S<sup>tri</sup> IgG + RBD<sub>v2</sub> IgG + NP<sub>v1</sub> IgG | — | — | — | 98.4%<br>(91.2%, 100.0%) | 95.9%<br>(83.9%, 99.6%) |
| S<sup>tri</sup> IgG + RBD<sub>v2</sub> IgG + NP<sub>v1</sub> IgG + S2 IgG | — | — | — | 98.9%<br>(86.1%, 100.0%) | 96.4%<br>(80.9%, 100.0%) |

**Supplementary Figure S1:**
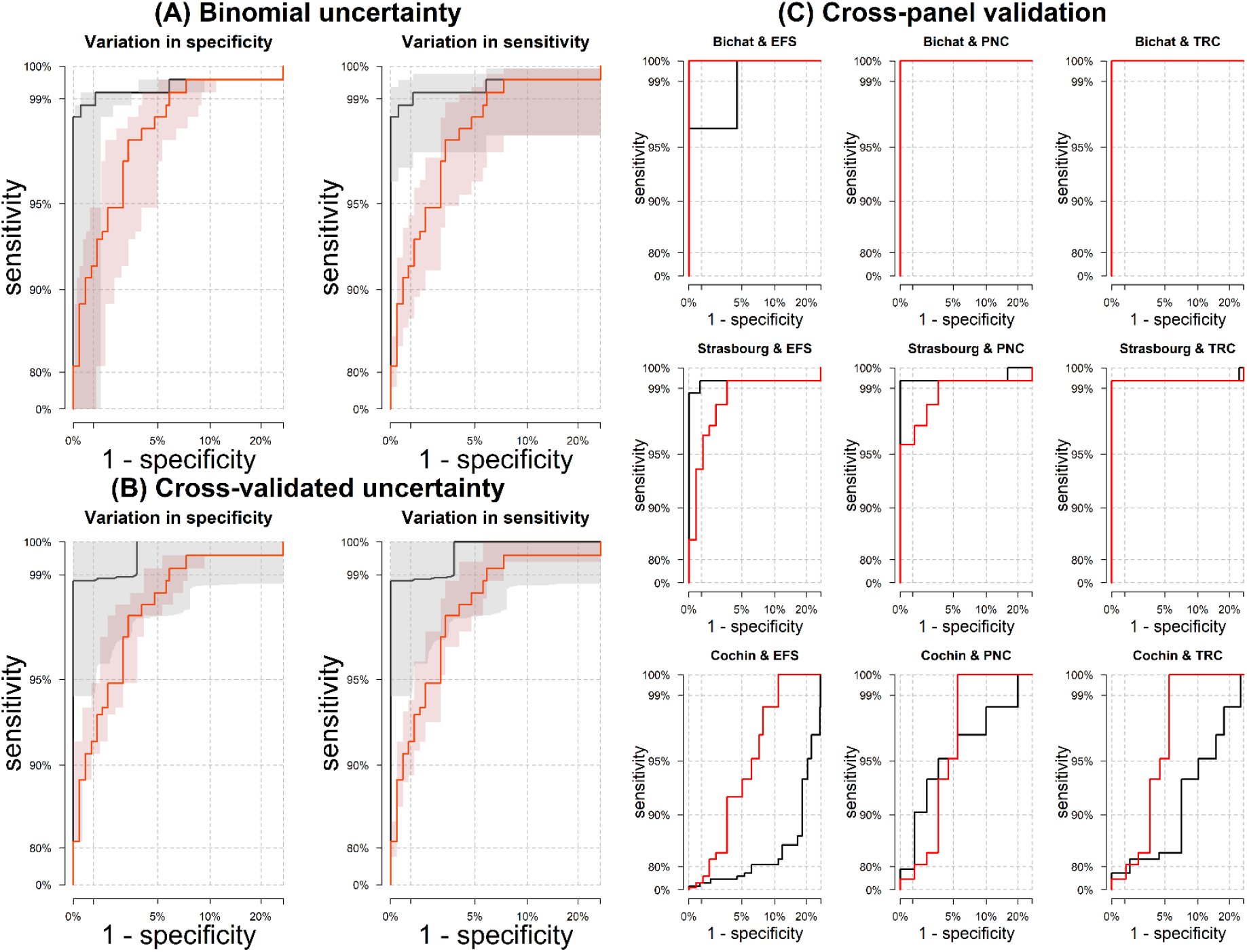
Quantification of uncertainty for serological classification. Results are shown for a single antigen (S^tri^) IgG assay in red, and a six antigen multiplex classifier in black. **(A)** Uncertainty estimated using Wilson’s binomial method applied to data from all samples. Uncertainty in specificity at fixed sensitivity, and variation in sensitivity at fixed specificity are shown separately. **(B)** Uncertainty estimated using 1000-fold cross-validation with training (2/3 of samples) and testing (1/3 of samples) data sets. Uncertainty in specificity at fixed sensitivity, and variation in sensitivity at fixed specificity are shown separately. **(C)** Cross-panel validation. The title of each plot denotes the panels that were used for testing, while the other panels were used for training.

## Appendix: Mathematical modelling of the duration of the anti-SARS-CoV-2 antibody response

### Overview

There are limited available longitudinal data on SARS-CoV-2 antibody kinetics, and no data from long-term follow-up (as of June 2020). However, there are a number of published studies on the long-term antibody kinetics to other coronaviruses, most notably Severe Acute Respiratory Syndrome coronavirus (SARS-CoV). Here we review some of the available published data, and describe how this can be used to provide prior information for modelling SARS-CoV-2 antibody kinetics.

### Prior longitudinal data on long-term antibody responses to coronaviruses

Appendix Table A1 summarises some of the published data on the long-term antibody kinetics to a number of coronaviruses: SARS-CoV, human seasonal coronavirus 229E, and Middle East Respiratory Syndrome coronavirus (MERS-CoV). From the extracted time series, we estimated two summary statistics characterizing the long-term antibody response: the half-life of the long-lived component of the antibody response, and the percentage reduction in antibody response after one year. The half-life of the long-lived component of the antibody response was estimated by fitting a linear regression model to measurements of (log) antibody response taken greater than six months after symptom onset. The percentage reduction in antibody response after one year was estimated based on the reduction from the peak measured antibody response to the estimated antibody level at one year. Although a wide range of assays from ELISA to micro-neutralisation were used in the reviewed studies, in this simple and approximate analysis we did not attempt to account for assay dependent effects, except to subtract background antibody levels where necessary.

Based on the estimated summary statistics, we assume that the long-term IgG antibody kinetics can be characterized as having a half-life of *d*_*l*_ = 400 days with a 60% reduction after one year. In terms of the parameters of the mathematical model of antibody kinetics, this corresponds to prior estimates of *c*_*l*_ = log(2)/*d*_*l*_ = 0.0017 and *ρ* ∼ 0.9. For sensitivity analyses, we also considered scenarios where *d*_l_ = 200 days and *d*_l_ = 800 days.

For IgM antibody kinetics, we assumed *d*_l_ = 100 days and *ρ* ∼ 0.9. For sensitivity analyses, we also considered scenarios where *d*_l_ = 50 days and *d*_l_ = 200 days.

**Appendix Table A1:**
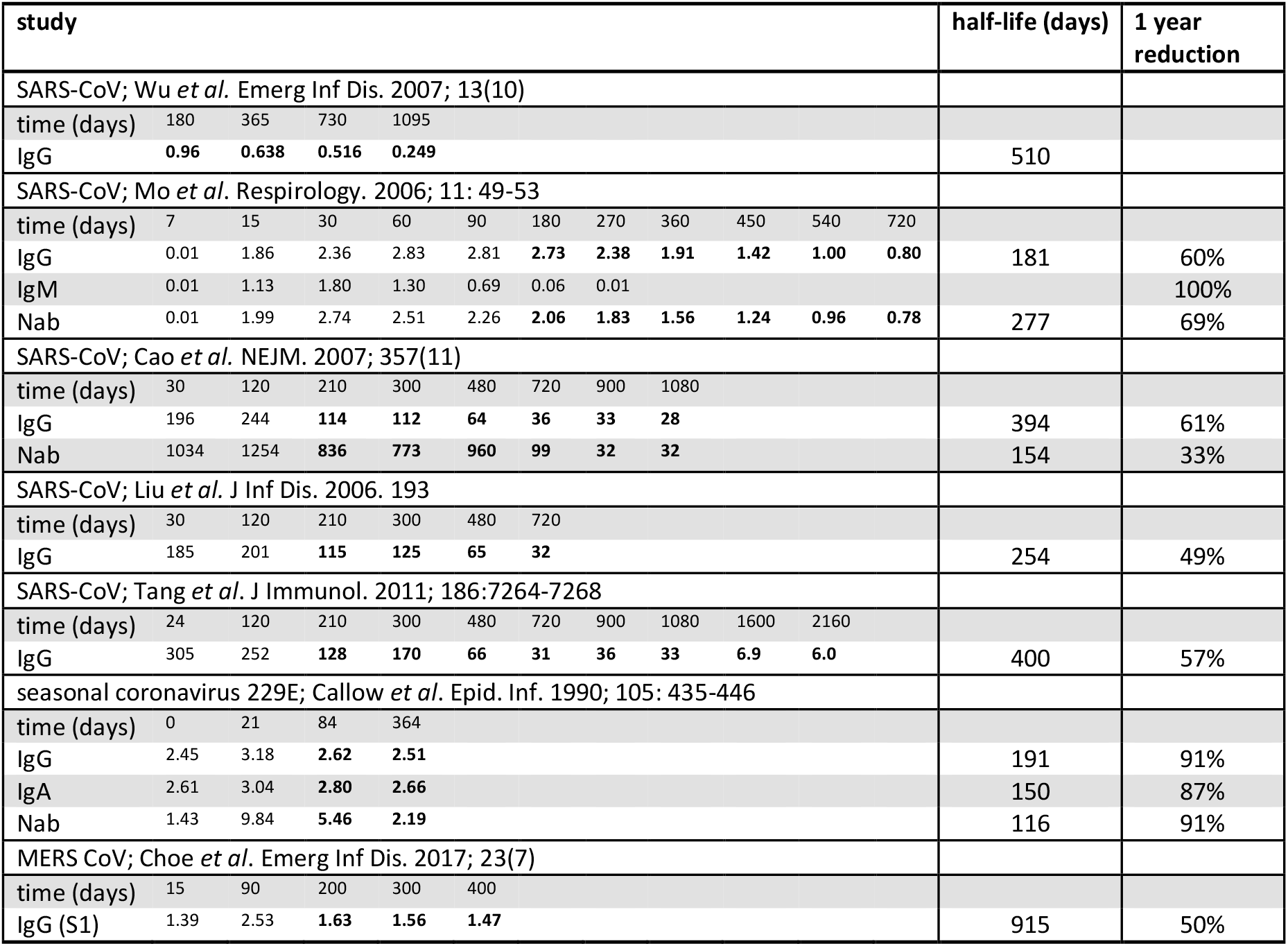
Prior data on the duration of antibody responses to coronaviruses. Data from longitudinal studies on measured antibody levels to SARS coronavirus, seasonal coronavirus 229E, and MERS coronavirus. For each study, the time series describing the antibody kinetics was extracted. The half-life of the long-lived component of the antibody response was estimated using measurements of antibody response measured after 6 months from symptom onset – the subset of the data used for this calculation is indicated in bold below. The percentage reduction in antibodies after one year is estimated based on the reduction from the peak measured response to the estimate antibody level at year.

### Case study of early-stage SARS-Cov-2 antibody kinetics: hospitalized patients in Hong Kong

We performed a secondary analysis of data from patients admitted to Princess Margaret Hospital and Queen Mary Hospital in Hong Kong, following the primary analysis by To, Tsang *et al* [1]. 23 patients with RT-qPCR confirmed SARS-CoV-2 infection were followed longitudinally for up to four weeks after initial onset of symptoms. Ten patients had severe COVID-19, all of whom required oxygen supplementation, and 13 patients had mild disease.

The Hong Kong based team expressed and purified recombinant proteins for receptor-binding domain (RBD) and nucleoprotein (NP). Genes encoding the spike RBD (amino acid residues 306 to 543 of the spike protein) and full length NP of SARS-CoV-2 were codon-optimized, synthesized and cloned. IgG and IgM antibody responses were quantified via the optical density (OD) from an enzyme immunoassay (EIA). Serial dilutions from 1:100 to 1:16,000 of a positive control serum were assayed for IgG responses. This allowed conversion of IgG antibody responses measured by EIA OD to dilutions. To determine the sero-positivity cutoff, the mean value of 93 anonymous archived serum specimens from 2018 plus 3 standard deviations was used. The cutoff values were: anti-NP IgG = 0.523 OD; anti-RBD IgG = 0.108 OD; anti-NP IgM = 0.177 OD; and anti-RBD IgM = 0.085. After conversion of the EIA OD values to dilutions, the sero-positivity cutoffs for IgG antibody responses were anti-NP IgG = 0.00682; and anti-RBD IgG = 0.002665.

### Results

Estimated model parameters are presented in Appendix Table A2. Appendix Figure A1 provides an overview of the fitted antibody kinetics to all participants. Detailed individual-level fits to the data, with quantification of uncertainty are shown in Appendix Figures A2-A5. Comparing the early kinetics of the IgG and IgM response, we estimate that the time to anti-NP IgG sero-conversion was 11.0 days (inter-quartile range (IQR): 8.1, 11.6), and the time to anti-NP IgM sero-conversion was 11.9 days (IQR: 8.4, 15.8). The time to anti-RBD IgG sero-conversion was 8.6 days (IQR: 5.3, 10.4), and the time to anti-NP IgM sero-conversion was 11.6 days (IQR: 9.2, 28.6). Although time to sero-conversion is dependent on the selection of sero-positivity cutoff, this suggests that IgM responses are not induced before IgG responses, and that both are generated at approximately the same time.

**Appendix Figure A1:**
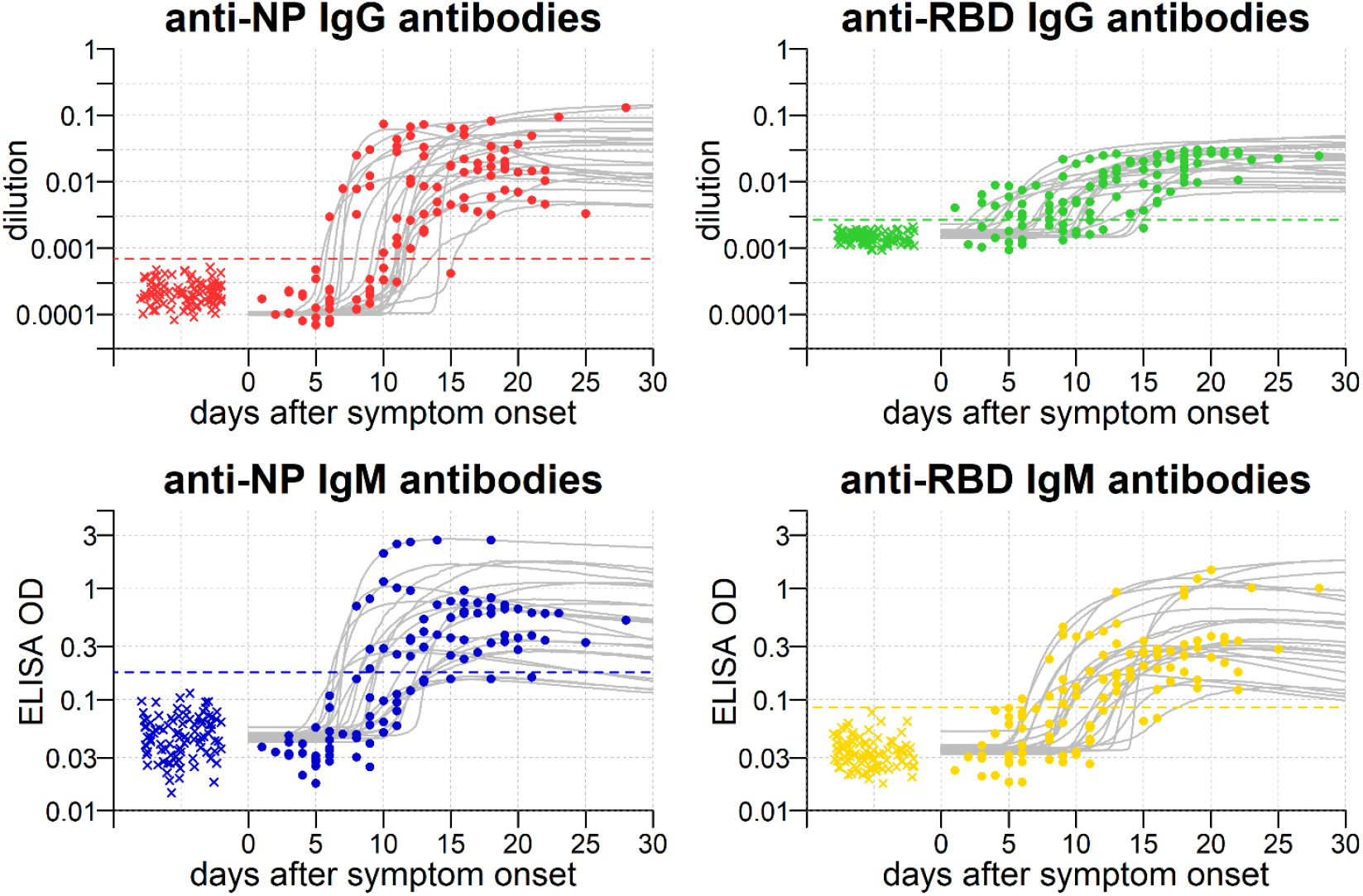
SARS-CoV-2 antibody kinetics in Hong Kong patients. Anti-nucleoprotein (NP) and anti-receptor-binding domain (RBD) antibody responses in 22 patients with PCR confirmed SARS-CoV-2 infection admitted to hospitals in Hong Kong. Measured antibody levels in patients are depicted as points. Measured antibody levels in negative controls are depicted as crosses. Grey lines show posterior median model prediction. The uncertainty of the model predictions is presented via 95% credible intervals in Figures s2-5. The horizontal dashed line represents the cutoff for sero-positivity.

**Appendix Figure A2:**
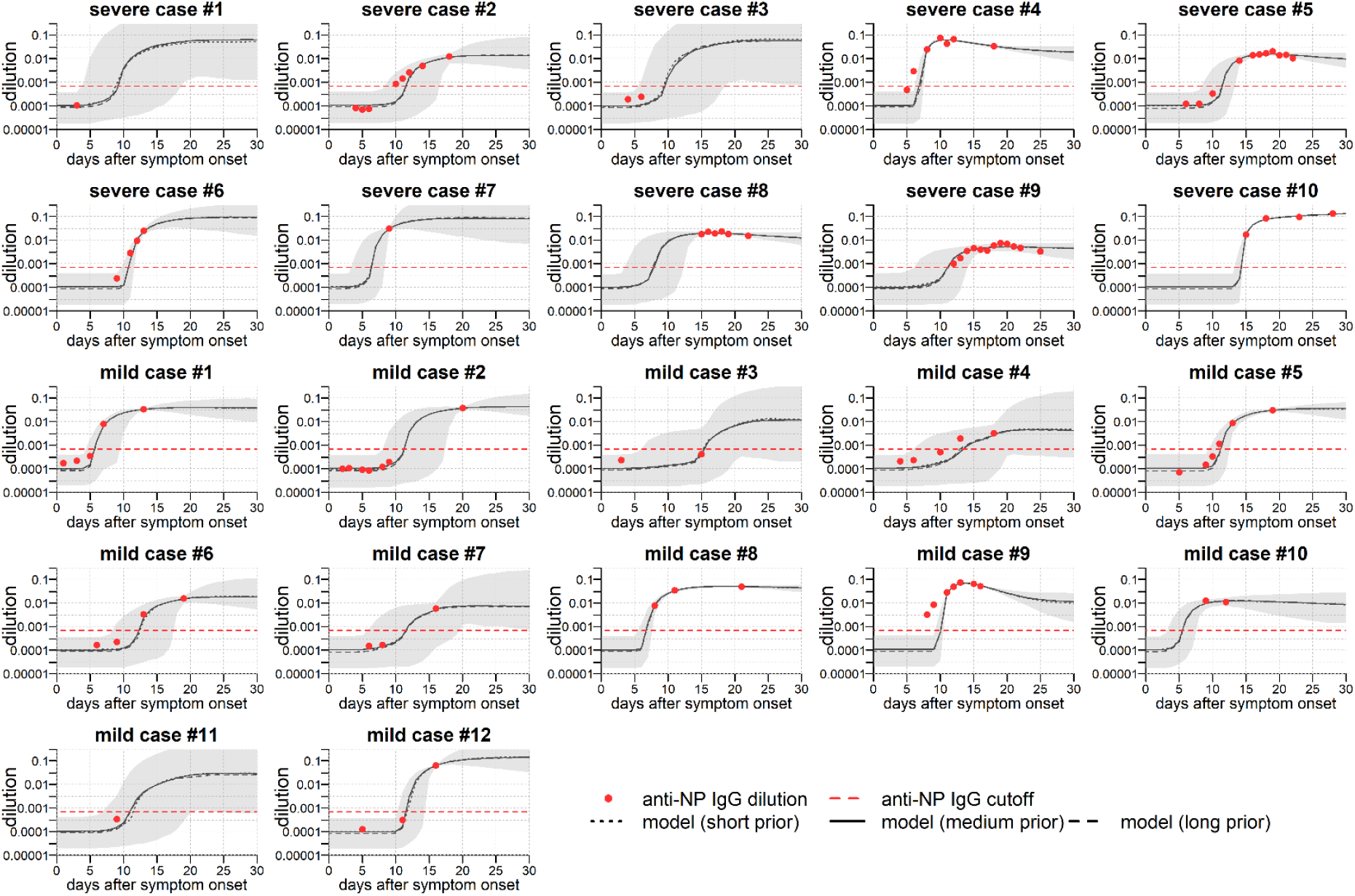
Model fit to short-term data on anti-NP IgG antibody responses. Measured antibody responses are shown as red points. Posterior median model predictions are shown as black lines, with 95% credible intervals in grey. The horizontal dashed line represents the cutoff for sero-positivity. Note that as there is no data on the long-term antibody response to SARS-CoV-2, three different sources of prior information were utilized. The half-life of the long-lived component of the antibody responses was assumed to be 200 days (short prior), 400 days (medium prior), or 800 days (long prior). Note that each of the three assumptions give near identical fits for the short-term kinetics displayed here.

**Appendix Figure A3:**
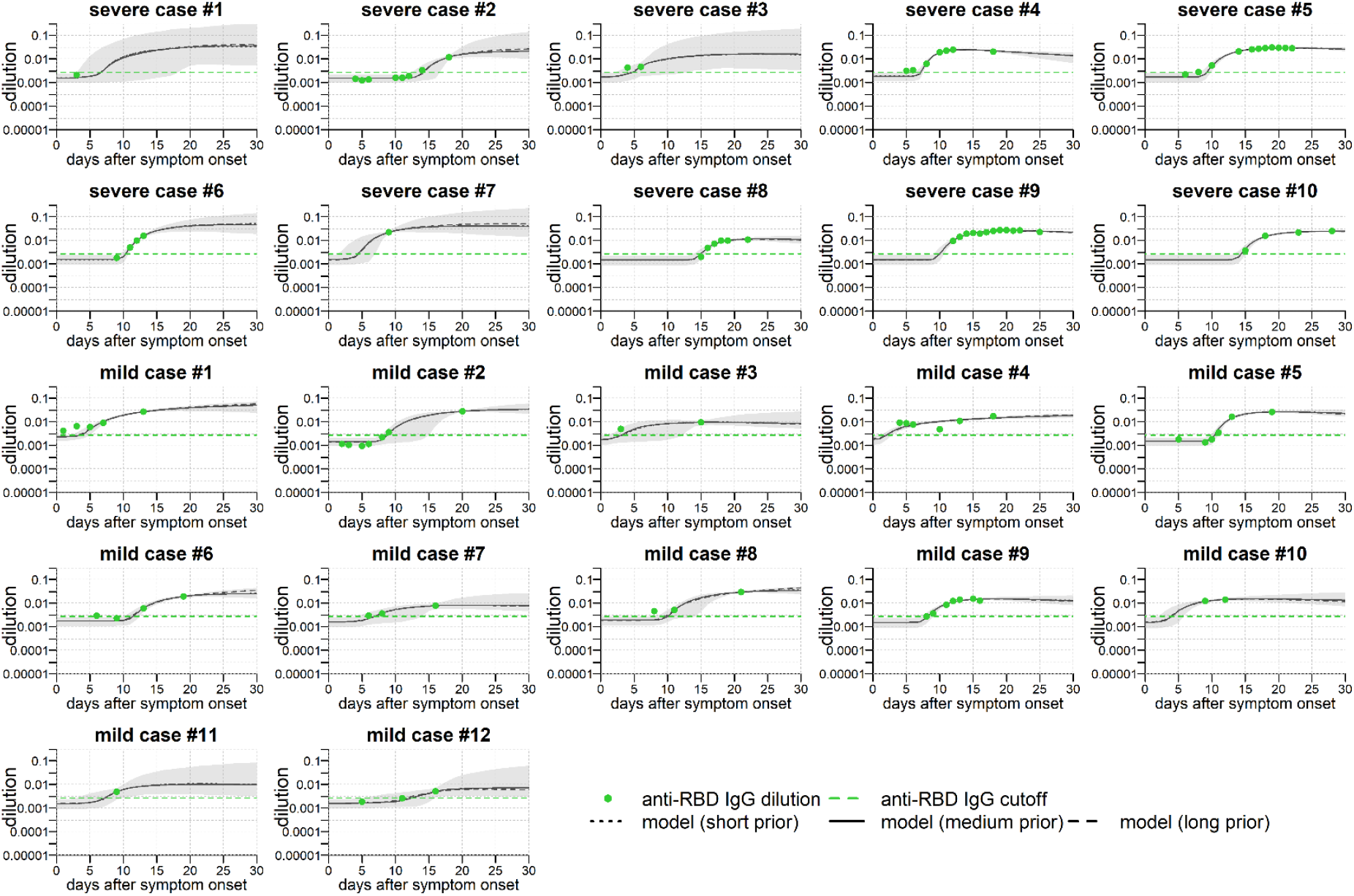
Model fit to short-term data on anti-RBD IgG antibody responses. Measured antibody responses are shown as red points. Posterior median model predictions are shown as black lines, with 95% credible intervals in grey. The horizontal dashed line represents the cutoff for sero-positivity. Note that as there is no data on the long-term antibody response to SARS-CoV-2, three different sources of prior information were utilized. The half-life of the long-lived component of the antibody responses was assumed to be 200 days (short prior), 400 days (medium prior), or 800 days (long prior). Note that each of the three assumptions give near identical fits for the short-term kinetics displayed here.

**Appendix Figure A4:**
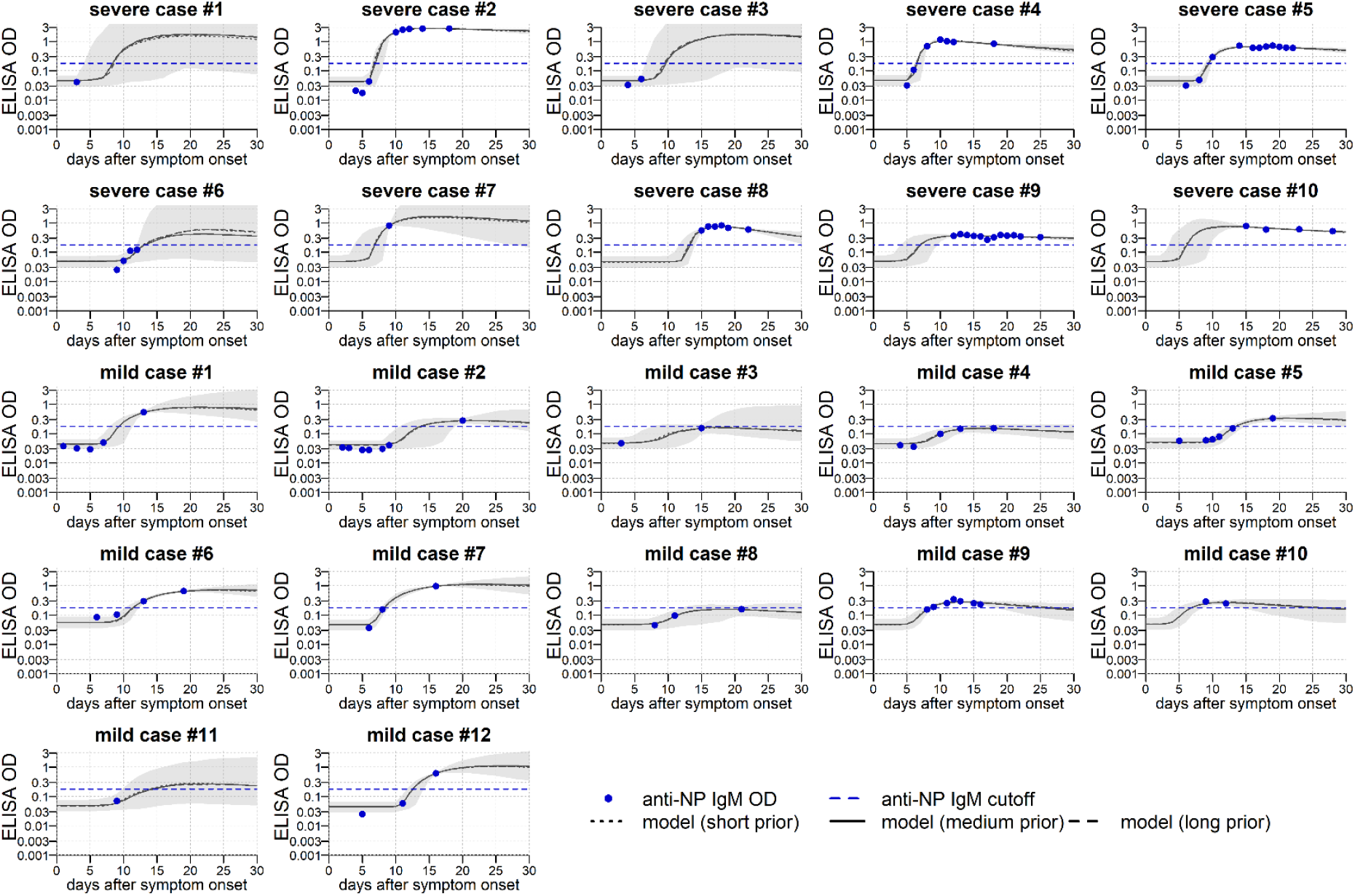
Model fit to short-term data on anti-NP IgM antibody responses. Measured antibody responses are shown as red points. Posterior median model predictions are shown as black lines, with 95% credible intervals in grey. The horizontal dashed line represents the cutoff for sero-positivity. Note that as there is no data on the long-term antibody response to SARS-CoV-2, three different sources of prior information were utilized. The half-life of the long-lived component of the antibody responses was assumed to be 50 days (short prior), 100 days (medium prior), or 200 days (long prior). Note that each of the three assumptions give near identical fits for the short-term kinetics displayed here.

**Appendix Figure A5:**
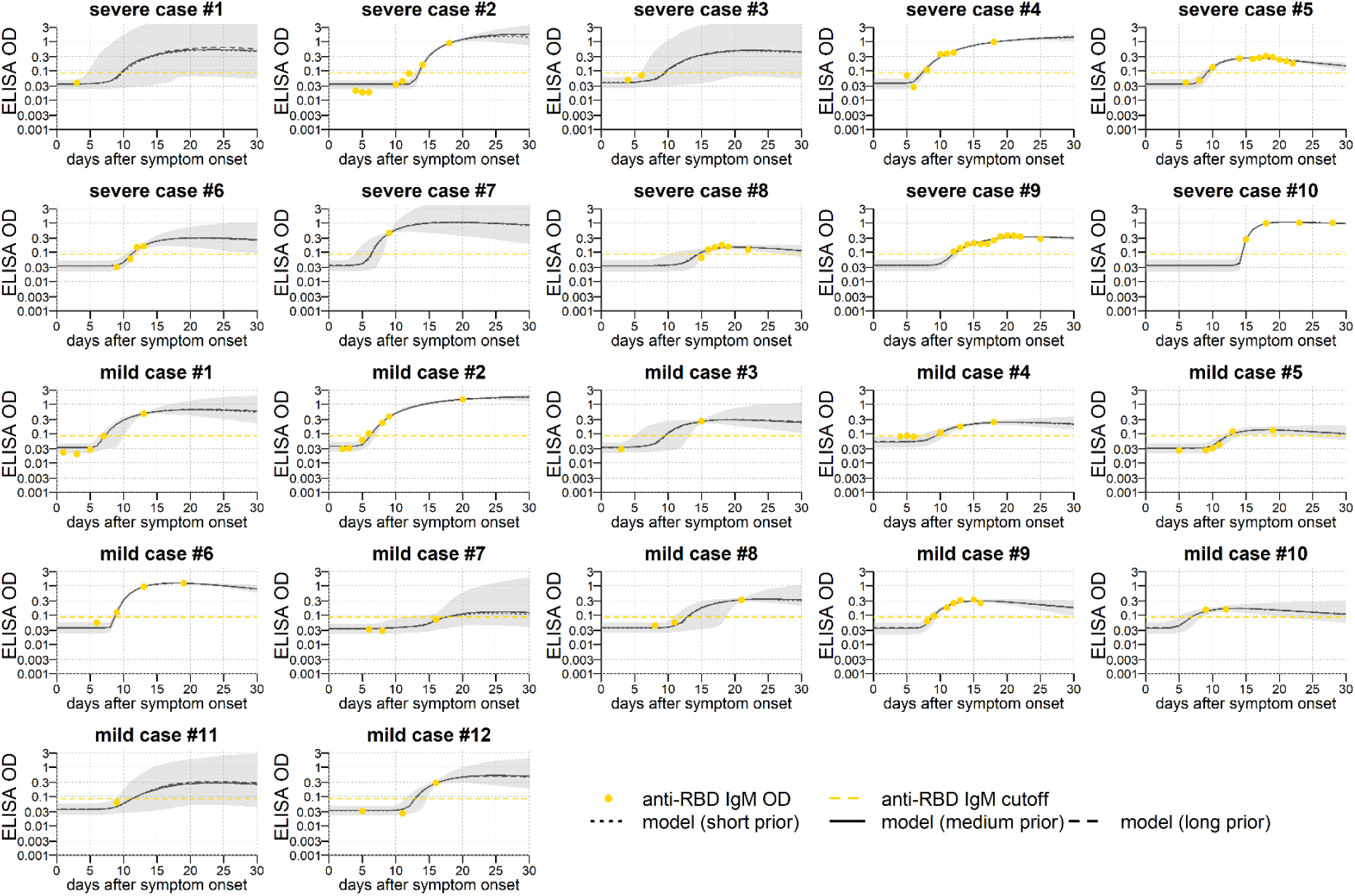
Model fit to short-term data on anti-RBD IgM antibody responses. Measured antibody responses are shown as red points. Posterior median model predictions are shown as black lines, with 95% credible intervals in grey. The horizontal dashed line represents the cutoff for sero-positivity. Note that as there is no data on the long-term antibody response to SARS-CoV-2, three different sources of prior information were utilized. The half-life of the long-lived component of the antibody responses was assumed to be 50 days (short prior), 100 days (medium prior), or 200 days (long prior). Note that each of the three assumptions give near identical fits for the short-term kinetics displayed here.

**Appendix Table A2:** Parameter estimates for antibody kinetics model fitted to Hong Kong data. Parameters of the antibody kinetics model are presented as posterior medians with 95% credible intervals. The model is fitted in a mixed-effects framework, so for every parameter we estimate the distribution within the entire population rather than a fixed value. We present the mean and standard deviation as summary statistics for the estimated distributions

| description | parameter | prior | NP IgG | RBD IgG | NP IgM | RBD IgM |
| --- | --- | --- | --- | --- | --- | --- |
| *mean of population-level distribution* |  |  |  |  |  |  |
| background IgG level | $A_{bg}$ | 0.001<br>($1.1 \times 10^{-6}$, 1.1) | 0.00011<br>($2.6 \times 10^{-5}$, 0.0003) | 0.0015<br>(0.0013, 0.0017) | — | — |
| background IgM level | $A_{bg}$ | 0.03<br>(0.001, 1.0) | — | — | 0.049<br>(0.043, 0.054) | 0.036<br>(0.032, 0.04) |
| ASC boost in mild cases (IgG) | $\beta_{mild}$ | 0.01<br>(0.0001, 1.2) | 0.014<br>(0.006, 0.051) | 0.0028<br>(0.0015, 0.0053) | — | — |
| ASC boost in mild cases (IgM) | $\beta_{mild}$ | 0.11<br>(0.01, 1.2) | — | — | 0.085<br>(0.048, 0.17) | 0.08<br>(0.04, 0.16) |
| ASC boost in severe cases (IgG) | $\beta_{sev}$ | 0.01<br>(0.0001, 1.2) | 0.028<br>(0.01, 0.207) | 0.0056<br>(0.0034, 0.0099) | — | — |
| ASC boost in severe cases (IgM) | $\beta_{sev}$ | 0.11<br>(0.01, 1.2) | — | — | 0.67<br>(0.29, 2.8) | 0.14<br>(0.07, 0.46) |
| delay in generation of antibody response (days) | $\tau$ | 5.4<br>(2.5, 15.1) | 9.6<br>(7.7, 11.9) | 7.8<br>(5.6, 11.7) | 7.9<br>(6.4, 9.8) | 8.7<br>(7.0, 10.7) |
| half-life of memory cells (days) | $d_m$ | 2.1<br>(1.5, 4.0) | 2.0<br>(1.3, 7.8) | 1.8<br>(1.3, 2.8) | 2.0<br>(1.3, 5.7) | 2.2<br>(1.5, 4.9) |
| half-life of short-lived ASCs (days) | $d_s$ | 3.2<br>(1.9, 9.2) | 2.5<br>(1.8, 4.1) | 2.4<br>(1.8, 3.7) | 2.4<br>(1.7, 3.8) | 2.8<br>(2.0, 4.7) |
| half-life of long-lived ASCs (days) (IgG) | $d_l$ | 400<br>(302, 567) | 408<br>(227, 727) | 417<br>(230, 771) | — | — |
| half-life of long-lived ASCs (days) (IgM) | $d_l$ | 100<br>(76, 142) | — | — | 104<br>(68, 163) | 103<br>(66, 167) |
| half-life of IgG molecules (days) | $d_a$ | 21<br>(18.7, 24.1) | 43.5<br>(25.7, 243.6) | 21.3<br>(18.4, 28.7) | — | — |
| half-life of IgM molecules (days) | $d_a$ | 10<br>(9.1, 11.5) | — | — | 10.8<br>(9.3, 164.2) | 10.2<br>(9.2, 13.2) |
| proportion of short-lived ASCs | $\rho$ | 90%<br>(65%, 95%) | 90%<br>(79%, 94%) | 80%<br>(57%, 94%) | 93%<br>(65%, 97%) | 89%<br>(62%, 98%) |
| *standard deviation of population-level distribution* |  |  |  |  |  |  |
| background IgG level | $A_{bg}$ | 0.0006<br>($6 \times 10^{-7}$, 0.8) | $5.7 \times 10^{-5}$<br>($1.0 \times 10^{-5}$, 0.00013) | 0.0004<br>(0.0003, 0.0005) | — | — |
| background IgM level | $A_{bg}$ | 0.01<br>(0.0003, 0.5) | — | — | 0.01<br>(0.007, 0.015) | 0.008<br>(0.006, 0.011) |
| ASC boost in mild cases (IgG) | $\beta_{mild}$ | 0.006<br>($5.4 \times 10^{-5}$, 0.9) | 0.020<br>(0.006, 0.23) | 0.0017<br>(0.0007, 0.005) | — | — |
| ASC boost in mild cases (IgM) | $\beta_{mild}$ | 0.06<br>(0.004, 1.1) | — | — | 0.045<br>(0.020, 0.17) | 0.06<br>(0.03, 0.21) |
| ASC boost in severe cases (IgG) | $\beta_{sev}$ | 0.006<br>($5.4 \times 10^{-5}$, 0.9) | 0.048<br>(0.01, 2.0) | 0.0030<br>(0.0015, 0.008) | — | — |
| ASC boost in severe cases (IgM) | $\beta_{sev}$ | 0.06<br>(0.004, 1.1) | — | — | 0.55<br>(0.19, 4.9) | 0.17<br>(0.06, 1.4) |
| delay in generation of antibody response (days) | $\tau$ | 3.5<br>(1.2, 34.6) | 4.2<br>(2.8, 6.9) | 6.5<br>(3.8, 17.5) | 3.5<br>(2.3, 5.9) | 3.8<br>(2.6, 6.4) |
| half-life of memory cells (days) | $d_m$ | 1.1<br>(0.5, 7.2) | 1.8<br>(0.6, 35.3) | 1.0<br>(0.5, 3.5) | 1.8<br>(0.6, 18.5) | 1.8<br>(0.7, 11.87) |
| half-life of short-lived ASCs (days) | $d_s$ | 2.3<br>(0.9, 29.2) | 1.3<br>(0.6, 3.5) | 1.2<br>(0.6, 2.8) | 1.2<br>(0.6, 3.1) | 1.6<br>(0.7, 4.6) |
| half-life of long-lived ASCs (days) (IgG) | $d_l$ | 109<br>(56, 349) | 111<br>(47, 384) | 114<br>(47, 404) | — | — |
| half-life of long-lived ASCs (days) (IgM) | $d_l$ | 22<br>(10, 69) | — | — | 22<br>(11, 67) | 23<br>(11, 73) |
| half-life of IgG molecules (days) | $d_a$ | 3.2<br>(1.8, 8.6) | 84.0<br>(22, 2808) | 5.4<br>(2.0, 27) | — | — |
| half-life of IgM molecules (days) | $d_a$ | 2.2<br>(1.2, 6.2) | — | — | 4.4<br>(1.5, 2770) | 2.6<br>(1.3, 12) |
| proportion of short-lived ASCs | $\rho$ | 0.07<br>(0.02, 0.40) | 0.06<br>(0.02, 0.26) | 0.25<br>(0.04, 0.45) | 0.08<br>(0.02, 0.42) | 0.18<br>(0.02, 0.44) |
| *observational variance* |  |  |  |  |  |  |
| standard deviation for ELISA measurements (IgG) | $\sigma_{obs}$ | 0.004<br>(0.0002, 0.1) | 0.0026<br>(0.0023, 0.0030) | 0.0011<br>(0.0009, 0.0013) | — | — |
| standard deviation for ELISA measurements (IgM) | $\sigma_{obs}$ | 0.04<br>(0.002, 1) | — | — | 0.031<br>(0.025, 0.037) | 0.022<br>(0.019, 0.025) |

**Appendix Table A3:**
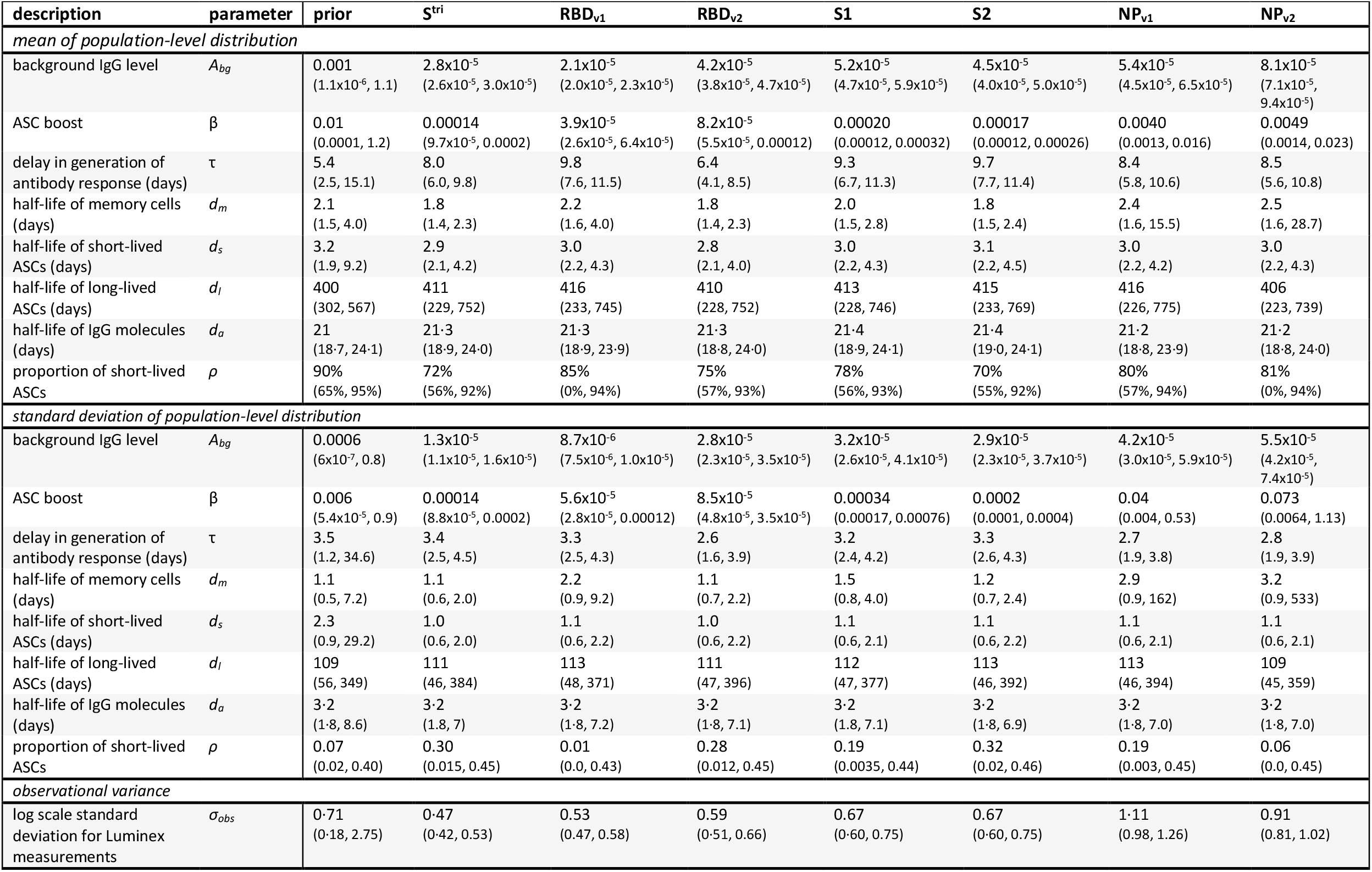
Parameter estimates for antibody kinetics model fitted to France data. Parameters of the antibody kinetics model are presented as posterior medians with 95% credible intervals. The model is fitted in a mixed-effects framework, so for every parameter we estimate the distribution within the entire population rather than a fixed value. We present the mean and standard deviation as summary statistics for the estimated distributions

| description | parameter | prior | S<sup>tri</sup> | RBD<sub>v1</sub> | RBD<sub>v2</sub> | S1 | S2 | NP<sub>v1</sub> | NP<sub>v2</sub> |
| --- | --- | --- | --- | --- | --- | --- | --- | --- | --- |
| *mean of population-level distribution* |  |  |  |  |  |  |  |  |  |
| background IgG level | $A_{bg}$ | 0.001<br>(1.1x10<sup>-6</sup>, 1.1) | 2.8x10<sup>-5</sup><br>(2.6x10<sup>-5</sup>, 3.0x10<sup>-5</sup>) | 2.1x10<sup>-5</sup><br>(2.0x10<sup>-5</sup>, 2.3x10<sup>-5</sup>) | 4.2x10<sup>-5</sup><br>(3.8x10<sup>-5</sup>, 4.7x10<sup>-5</sup>) | 5.2x10<sup>-5</sup><br>(4.7x10<sup>-5</sup>, 5.9x10<sup>-5</sup>) | 4.5x10<sup>-5</sup><br>(4.0x10<sup>-5</sup>, 5.0x10<sup>-5</sup>) | 5.4x10<sup>-5</sup><br>(4.5x10<sup>-5</sup>, 6.5x10<sup>-5</sup>) | 8.1x10<sup>-5</sup><br>(7.1x10<sup>-5</sup>, 9.4x10<sup>-5</sup>) |
| ASC boost | $\beta$ | 0.01<br>(0.0001, 1.2) | 0.00014<br>(9.7x10<sup>-5</sup>, 0.0002) | 3.9x10<sup>-5</sup><br>(2.6x10<sup>-5</sup>, 6.4x10<sup>-5</sup>) | 8.2x10<sup>-5</sup><br>(5.5x10<sup>-5</sup>, 0.00012) | 0.00020<br>(0.00012, 0.00032) | 0.00017<br>(0.00012, 0.00026) | 0.0040<br>(0.0013, 0.016) | 0.0049<br>(0.0014, 0.023) |
| delay in generation of antibody response (days) | $\tau$ | 5.4<br>(2.5, 15.1) | 8.0<br>(6.0, 9.8) | 9.8<br>(7.6, 11.5) | 6.4<br>(4.1, 8.5) | 9.3<br>(6.7, 11.3) | 9.7<br>(7.7, 11.4) | 8.4<br>(5.8, 10.6) | 8.5<br>(5.6, 10.8) |
| half-life of memory cells (days) | $d_m$ | 2.1<br>(1.5, 4.0) | 1.8<br>(1.4, 2.3) | 2.2<br>(1.6, 4.0) | 1.8<br>(1.4, 2.3) | 2.0<br>(1.5, 2.8) | 1.8<br>(1.5, 2.4) | 2.4<br>(1.6, 15.5) | 2.5<br>(1.6, 28.7) |
| half-life of short-lived ASCs (days) | $d_s$ | 3.2<br>(1.9, 9.2) | 2.9<br>(2.1, 4.2) | 3.0<br>(2.2, 4.3) | 2.8<br>(2.1, 4.0) | 3.0<br>(2.2, 4.3) | 3.1<br>(2.2, 4.5) | 3.0<br>(2.2, 4.2) | 3.0<br>(2.2, 4.3) |
| half-life of long-lived ASCs (days) | $d_l$ | 400<br>(302, 567) | 411<br>(229, 752) | 416<br>(233, 745) | 410<br>(228, 752) | 413<br>(228, 746) | 415<br>(233, 769) | 416<br>(226, 775) | 406<br>(223, 739) |
| half-life of IgG molecules (days) | $d_a$ | 21<br>(18.7, 24.1) | 21.3<br>(18.9, 24.0) | 21.3<br>(18.9, 23.9) | 21.3<br>(18.8, 24.0) | 21.4<br>(18.9, 24.1) | 21.4<br>(19.0, 24.1) | 21.2<br>(18.8, 23.9) | 21.2<br>(18.8, 24.0) |
| proportion of short-lived ASCs | $\rho$ | 90%<br>(65%, 95%) | 72%<br>(56%, 92%) | 85%<br>(0%, 94%) | 75%<br>(57%, 93%) | 78%<br>(56%, 93%) | 70%<br>(55%, 92%) | 80%<br>(57%, 94%) | 81%<br>(0%, 94%) |
| *standard deviation of population-level distribution* |  |  |  |  |  |  |  |  |  |
| background IgG level | $A_{bg}$ | 0.0006<br>(6x10<sup>-7</sup>, 0.8) | 1.3x10<sup>-5</sup><br>(1.1x10<sup>-5</sup>, 1.6x10<sup>-5</sup>) | 8.7x10<sup>-6</sup><br>(7.5x10<sup>-6</sup>, 1.0x10<sup>-5</sup>) | 2.8x10<sup>-5</sup><br>(2.3x10<sup>-5</sup>, 3.5x10<sup>-5</sup>) | 3.2x10<sup>-5</sup><br>(2.6x10<sup>-5</sup>, 4.1x10<sup>-5</sup>) | 2.9x10<sup>-5</sup><br>(2.3x10<sup>-5</sup>, 3.7x10<sup>-5</sup>) | 4.2x10<sup>-5</sup><br>(3.0x10<sup>-5</sup>, 5.9x10<sup>-5</sup>) | 5.5x10<sup>-5</sup><br>(4.2x10<sup>-5</sup>, 7.4x10<sup>-5</sup>) |
| ASC boost | $\beta$ | 0.006<br>(5.4x10<sup>-5</sup>, 0.9) | 0.00014<br>(8.8x10<sup>-5</sup>, 0.0002) | 5.6x10<sup>-5</sup><br>(2.8x10<sup>-5</sup>, 0.00012) | 8.5x10<sup>-5</sup><br>(4.8x10<sup>-5</sup>, 3.5x10<sup>-5</sup>) | 0.00034<br>(0.00017, 0.00076) | 0.0002<br>(0.0001, 0.0004) | 0.04<br>(0.004, 0.53) | 0.073<br>(0.0064, 1.13) |
| delay in generation of antibody response (days) | $\tau$ | 3.5<br>(1.2, 34.6) | 3.4<br>(2.5, 4.5) | 3.3<br>(2.5, 4.3) | 2.6<br>(1.6, 3.9) | 3.2<br>(2.4, 4.2) | 3.3<br>(2.6, 4.3) | 2.7<br>(1.9, 3.8) | 2.8<br>(1.9, 3.9) |
| half-life of memory cells (days) | $d_m$ | 1.1<br>(0.5, 7.2) | 1.1<br>(0.6, 2.0) | 2.2<br>(0.9, 9.2) | 1.1<br>(0.7, 2.2) | 1.5<br>(0.8, 4.0) | 1.2<br>(0.7, 2.4) | 2.9<br>(0.9, 162) | 3.2<br>(0.9, 533) |
| half-life of short-lived ASCs (days) | $d_s$ | 2.3<br>(0.9, 29.2) | 1.0<br>(0.6, 2.0) | 1.1<br>(0.6, 2.2) | 1.0<br>(0.6, 2.2) | 1.1<br>(0.6, 2.1) | 1.1<br>(0.6, 2.2) | 1.1<br>(0.6, 2.1) | 1.1<br>(0.6, 2.1) |
| half-life of long-lived ASCs (days) | $d_l$ | 109<br>(56, 349) | 111<br>(46, 384) | 113<br>(48, 371) | 111<br>(47, 396) | 112<br>(47, 377) | 113<br>(46, 392) | 113<br>(46, 394) | 109<br>(45, 359) |
| half-life of IgG molecules (days) | $d_a$ | 3.2<br>(1.8, 8.6) | 3.2<br>(1.8, 7) | 3.2<br>(1.8, 7.2) | 3.2<br>(1.8, 7.1) | 3.2<br>(1.8, 7.1) | 3.2<br>(1.8, 6.9) | 3.2<br>(1.8, 7.0) | 3.2<br>(1.8, 7.0) |
| proportion of short-lived ASCs | $\rho$ | 0.07<br>(0.02, 0.40) | 0.30<br>(0.015, 0.45) | 0.01<br>(0.0, 0.43) | 0.28<br>(0.012, 0.45) | 0.19<br>(0.0035, 0.44) | 0.32<br>(0.02, 0.46) | 0.19<br>(0.003, 0.45) | 0.06<br>(0.0, 0.45) |
| *observational variance* |  |  |  |  |  |  |  |  |  |
| log scale standard deviation for Luminex measurements | $\sigma_{obs}$ | 0.71<br>(0.18, 2.75) | 0.47<br>(0.42, 0.53) | 0.53<br>(0.47, 0.58) | 0.59<br>(0.51, 0.66) | 0.67<br>(0.60, 0.75) | 0.67<br>(0.60, 0.75) | 1.11<br>(0.98, 1.26) | 0.91<br>(0.81, 1.02) |

